# Modeling the early phase of the Belgian COVID-19 epidemic using a stochastic compartmental model and studying its implied future trajectories

**DOI:** 10.1101/2020.06.29.20142851

**Authors:** Steven Abrams, James Wambua, Eva Santermans, Lander Willem, Elise Kuylen, Pietro Coletti, Pieter Libin, Christel Faes, Oana Petrof, Sereina A. Herzog, Philippe Beutels, Niel Hens

## Abstract

Following the onset of the ongoing COVID-19 pandemic throughout the world, a large fraction of the global population is or has been under strict measures of physical distancing and quarantine, with many countries being in partial or full lockdown. These measures are imposed in order to reduce the spread of the disease and to lift the pressure on healthcare systems. Estimating the impact of such interventions as well as monitoring the gradual relaxing of these stringent measures is quintessential to understand how resurgence of the COVID-19 epidemic can be controlled for in the future. In this paper we use a stochastic age-structured discrete time compartmental model to describe the transmission of COVID-19 in Belgium. Our model explicitly accounts for age-structure by integrating data on social contacts to (i) assess the impact of the lockdown as implemented on March 13, 2020 on the number of new hospitalizations in Belgium; (ii) conduct a scenario analysis estimating the impact of possible exit strategies on potential future COVID-19 waves. More specifically, the aforementioned model is fitted to hospital admission data, data on the daily number of COVID-19 deaths and serial serological survey data informing the (sero)prevalence of the disease in the population while relying on a Bayesian MCMC approach. Our age-structured stochastic model describes the observed outbreak data well, both in terms of hospitalizations as well as COVID-19 related deaths in the Belgian population. Despite an extensive exploration of various projections for the future course of the epidemic, based on the impact of adherence to measures of physical distancing and a potential increase in contacts as a result of the relaxation of the stringent lockdown measures, a lot of uncertainty remains about the evolution of the epidemic in the next months.

## 1 Introduction

The COVID-19 pandemic is caused by severe acute respiratory syndrome coronavirus 2 (SARS-CoV-2), a pathogenic infectious agent, which was initially identified in Wuhan (China), where several patients presented with pneumonia after developing symptoms between December 8, 2019 and January 2 [1]. COVID-19 was officially declared a pandemic by the WHO on March 11, 2020. More than 6 million confirmed cases and more than 380,000 deaths were reported globally by June 1, 2020, of which 58,000 confirmed cases and 9,500 deaths occurred in Belgium [2].

In line with other EU countries, the Belgian government issued a travel notice advising against non-essential flights to China, excluding Hong Kong, on January 29. As of March 6, a travel ban was issued for school trips to Italy. On March 10, Belgian authorities advised to cancel all indoor events of 1,000 participants or more. Furthermore, physical distancing measures were taken with companies being advised to allow their employees to work from home as much as possible. A closure of all schools, cafes and restaurants was ordered as well as a cancellation of all public gatherings as of March 13 at midnight. On March 17, the Belgian National Security Council announced additional measures to be taken, thereby imposing stricter measures of physical distancing, prohibiting non-essential travel to foreign countries and within the own borders (i.e., only allowing people to leave their homes to buy food or to go to work, at least when working in healthcare, transport or other essential professions), closure of shops providing non-essential services with the addition of penalties for everyone not abiding with the rules. A lockdown was imposed on Wednesday March 18 at noon. The borders were closed as of March 20. Throughout the epidemic, the government continuously stressed the importance of measures of physical distancing and hygiene, thereby avoiding physical contacts, ensuring regular washing of the hands, coughing and sneezing in the inner elbow, not shaking hands and staying at home when having COVID-19 related symptoms [3].

Upon having imposed very strict measures of physical distancing, including mobility restrictions and school closure, a thorough investigation of different exit strategies is required to relax unsustainable social life and economic constraints while maintaining control over pressure exerted on the health care system. After a specific exit strategy is implemented, a careful monitoring of the outbreak is necessary to avoid subsequent waves of COVID-19 infections.

Here we use a stochastic, discrete, age-structured compartmental model for COVID-19 transmission. The model is contrasted to Belgian data on the daily number of new hospitalizations and deaths prior and after mitigation strategies have been imposed. The model accounts for pre-symptomatic and asymptomatic transmission. Age-specific data on social contacts is used to inform transmission parameters [4] and serial serological survey data is incorporated in the model to inform the prevalence of past exposure to the disease [5]. The impact of the intervention measures as well as various exit strategies upon relaxing lockdown measures are studied in the context of this model.

The paper is organized as follows. In Section 2, we provide specific details on the (stochastic) compartmental model, its parametrization and estimation of model parameters based on several data sources. Moreover, we study the impact of intervention measures and subsequent exit strategies on the spread of COVID-19. The results of fitting the stochastic model to the available data is presented in Section 3. Furthermore, the impact of various exit strategies on the number of new hospitalizations is visualized using an extensive scenario-analysis. Finally, in Section 4, we discuss limitations and strengths of the proposed approach and we present avenues for further research.

## 2 Methodology

### 2.1 Mathematical compartmental transmission model

We use an adapted version of an SEIR mathematical compartmental model to describe COVID-19 disease dynamics. In this model, individuals are susceptible to infection when in compartment *S*, and after an effective contact (between a susceptible and infectious individual) the susceptible individual moves to an exposed state *E* at age- and time-specific rate ***λ***(*t*), referred to as the force of infection (with boldface notation representing a vector including age-specific rates). After a latent period, the individual becomes infectious and moves to a pre-symptomatic state *I*_*presym*_ at rate *γ*. Afterwards, individuals either develop symptoms (state *I*_*mild*_) with probability 1 − ***p*** or remain completely free of symptoms (compartment *I*_*asym*_, probability ***p***). Asymptomatic cases recover at rate *δ*_1_. Symptomatic infections are either very mild and such cases recover at rate 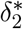 (for an age-dependent fraction of these individuals, represented by the vector ***ϕ***_0_ = ***δ***_2_*/*(***δ***_2_ + ***ψ***), where 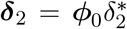 and 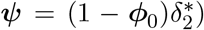 or they move to a state *I*_*sev*_ prior to requiring hospitalization (i.e., severe infection is defined as requiring hospitalization). When severely ill, implying hospitalization, individuals move to state *I*_*hosp*_ with probability ***ϕ***_1_, or become critically ill (*I*_*ICU*_) with probability 1 −***ϕ***_1_. Hospitalized and critically ill patients admitted to the Intensive Care Unit (ICU) recover at rate ***δ***_3_ and ***δ***_4_ with probabilities *{*1 − *µ*_*hosp*_(*a*)*}* and *{*1 − *µ*_*icu*_(*a*)*}*, respectively, where *µ*_*hosp*_(*a*) = ***τ*** _1_*/*(***τ*** _1_ + ***δ***_3_) and *µ*_*icu*_(*a*) = ***τ*** _2_*/*(***τ*** _2_ + ***δ***_4_) represent the age-specific case-fatality rates (i.e., probabilities of dying when severely ill and hospitalized on a general ward or admitted to ICU). Hospitalized and ICU patients die at rate ***τ*** _1_ or ***τ*** _2_ with probabilities *µ*_*hosp*_(*a*) and *µ*_*icu*_(*a*), respectively. A schematic overview of the compartmental model is given in Figure 1. Individuals in the red compartments are able to transmit the disease.

**Figure 1:**
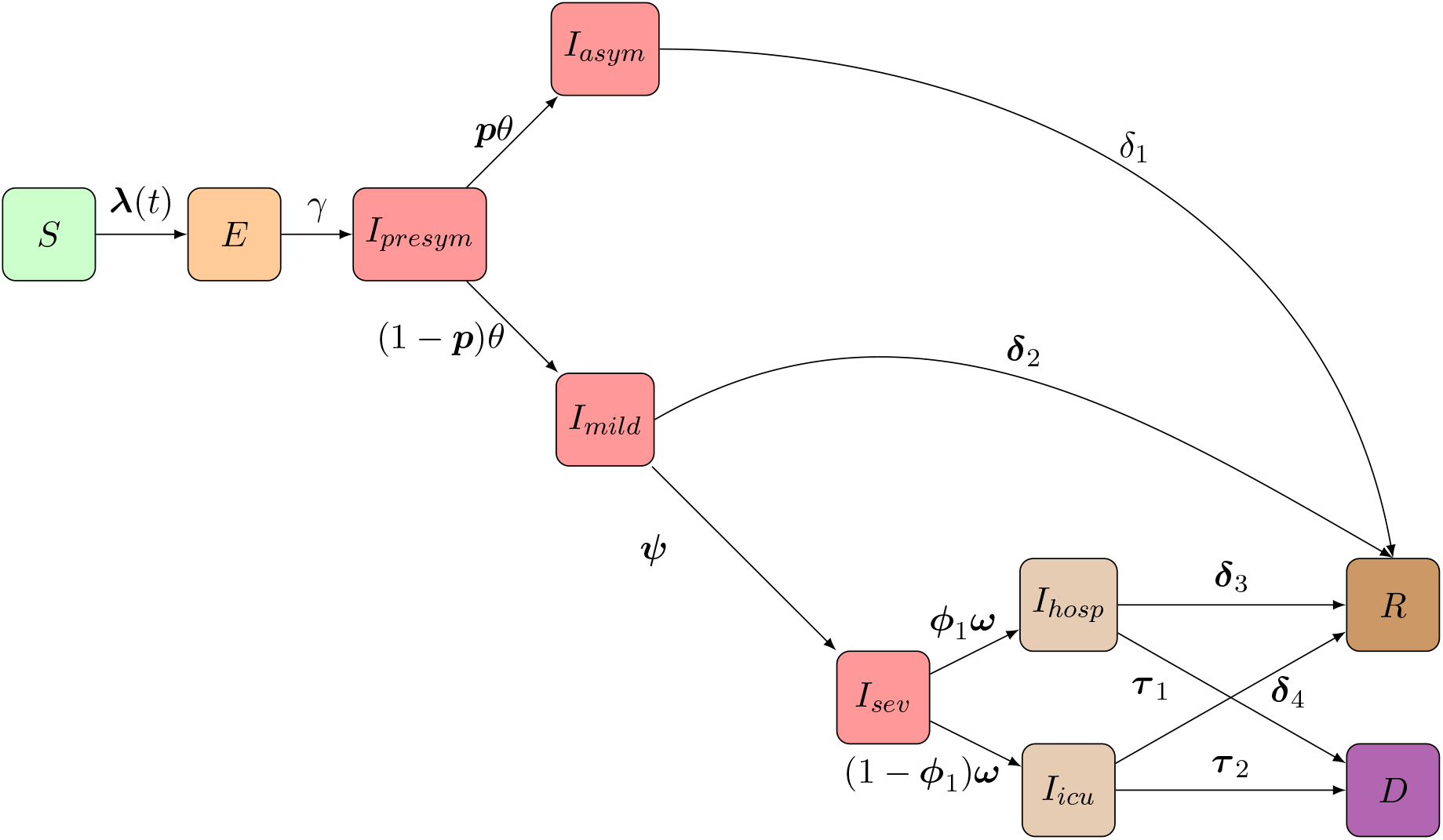
Schematic overview of the flows of individuals in the compartmental model: Following SARS-CoV-2/COVID-19 infection susceptible individuals (*S*) move to an exposed state (*E*) and after a latent period individuals further progress to a pre-symptomatic state (*I*_*presym*_) in which they can infect others. Consequently, individuals stay either completely symptom-free (*I*_*asym*_) or develop mild symptoms (*I*_*mild*_). Asymptomatic individuals will recover over time. Upon having mild symptoms, persons either recover (*R*) or require hospitalization (going from *I*_*sev*_ to *I*_*hosp*_ or *I*_*icu*_) prior to recovery (*R*) or death (*D*).

The following set of ordinary differential equations describes the flows in the (deterministic version of the) proposed age-structured compartmental model:

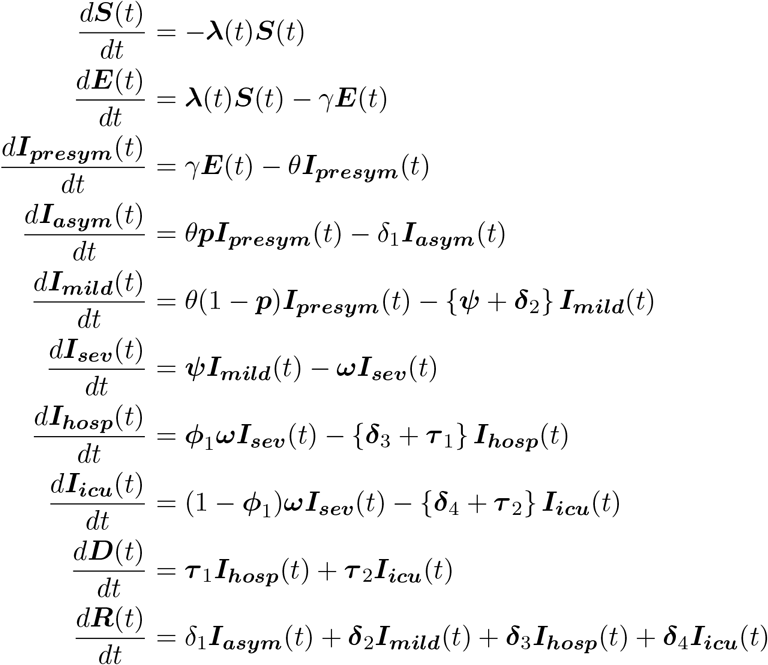

where, for example, ***S*** = (*S*_1_(*t*), *S*_2_(*t*), …, *S*_*K*_(*t*))^*T*^ represents the vector of number of susceptible individuals in age group *k* = 1, …, *K* in the population at time *t*. A full account on the definition of the different compartments and the notation used for the number of individuals therein can be found in Table A1 in Appendix A. An overview of the different parameter definitions can be found in Table B3.

The proposed age-structured compartmental transmission model consists of 10 age classes, i.e., [0-10), [10-20), [20-30), [30-40), [40-50), [50-60), [60-70), [70-80), [80-90), [90, ∞) with the number of individuals in each age class obtained from Eurostat.

### 2.2 Social contact data and transmission rates

As mentioned previously, the infectious phase of COVID-19 disease is divided into two different states: a pre-symptomatic state occurring before the end of the incubation period, followed by a state in which individuals may either remain asymptomatic or develop (mild to severe) symptoms (see Figure 1). Transmission of the disease is governed by an age- and time-dependent force of infection. The age-specific force of infection in age group *k* = 1, …, *K*, denoted by *λ*(*k, t*), is defined as the instantaneous rate at which a susceptible person in age group *k* acquires infection at time *t*. Furthermore, the time-invariant transmission rate *β*(*k, k* ′) represents the average per capita rate at which an infectious individual in age group *k* makes an effective contact with a susceptible individual in age group *k*, per unit of time. Consequently, the force of infection is defined as

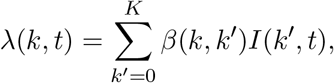

where *I*(*k* ′, *t*) denotes the total number of infectious individuals in age group *k* ′at time *t* and *β*(*k, k*′) can be rendered as

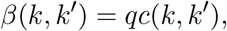

when relying on the so-called social contact hypothesis [6]. This hypothesis entails that *c*(*k, k* ′) are the per capita rates at which an individual in age group *k* makes contact with an individual in age group *k* ′, per unit of time, and *q* is a proportionality factor capturing contextual and host- and disease-specific characteristics such as susceptibility and infectiousness. The (*K* × *K*)-matrix *C* containing the elements *c*(*k, k* ′) is referred to as the social contact matrix describing mixing behaviour within and between different age groups in the population. Social contact rates *c*(*k, k*) are estimated based on social contact data from Flanders (Belgium) collected in 2010 [7, 8, 4, 9]. We hereby assume that contact rates for Flanders can be used for all regions in Belgium.

In this manuscript, we rely on social contact matrices *C*_*sym*_ and *C*_*asym*_ estimated for symptomatic and asymptomatic individuals, implying

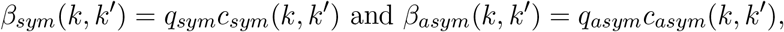

defining transmission rates for both symptomatic and asymptomatic cases, respectively (see Appendix C). Here, we assume that the relative infectiousness of symptomatic versus asymptomatic cases is equal to 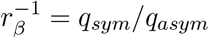. The age-dependent force of infection is defined as:

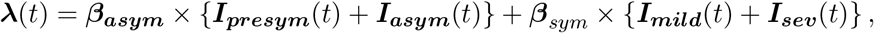

where ***λ***(*t*) = (*λ*(1, *t*), *λ*(2, *t*), …, *λ*(*K, t*)) written as a matrix multiplication with boldface notation for vectors and matrices. Note that hospitalized individuals are assumed not to contribute to the transmission process because of isolation.

### 2.3 Discrete time stochastic epidemic model

The spread of the virus is hampered by reductions in the number of contacts and changes in the way contacts are made, either voluntarily or as a consequence of government intervention. These time- (and age-) dependent behavioural changes introduce substantial uncertainty in the further course of the outbreak and require stochastic model components to evaluate the effectiveness of the intervention strategies and to make future predictions in terms of, for example, new hospitalizations. Moreover, stochastic epidemic models allow to determine the probability of extinction based on multiple realizations of the model. Therefore, we amended the deterministic model hitherto described into a discrete time stochastic epidemic model to describe the transmission process under the mitigation strategies as highlighted hereabove.

Our chain binomial model, originally introduced by Bailey [10], is a so-called discrete-time stochastic alternative to the continuous-time deterministic model based on the health states and transitions presented in Figure 1. The chain binomial model assumes a stochastic version of an epidemic obtained through a succession of discrete generations of infected individuals in a probabilistic manner. Consider a time interval (*t, t*+*h*], where *h* represents the length between two consecutive time points at which we evaluate the model, here *h* = 1*/*24 day. Let us assume that there are *S*_*t*_(*k*) susceptible individuals at time *t* in age group *k*, we expect 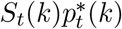 newly exposed individuals at time *t* + *h*, i.e.,

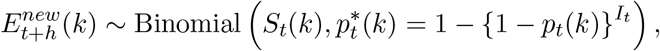

where *I*_*t*_ is the total number of infected individuals at time *t* and *p*_*t*_(*k*) represents the transmission probability conditional upon contact between a susceptible individual in age group *k* and an infected individual. The probability that a susceptible individual escapes infection (during a single contact with an infected individual) is equal to *q*_*t*_(*k*) = 1 − *p*_*t*_(*k*), hence, assuming all contacts to be equally infectious, the escape probability is 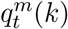 in case the susceptible individual contacts *m* infectious individuals. In this setting, the probability of infection 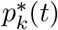 for a susceptible individual in age group *k* = 1, … *K* can be obtained as:

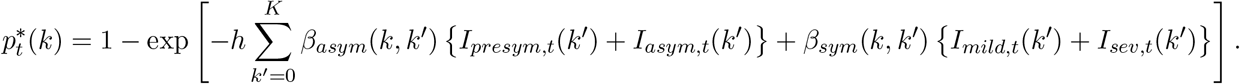

The number of individuals in age group *k* leaving the exposed state (and entering the pre-symptomatic compartment) within the specified time interval is

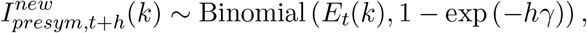

where 1*/γ* equals the mean length of the latency period. Probabilistic transitions in the other compartments are derived similarly, hence, a discretized age-structured stochastic model (with step size *h* = 1*/*24 days) is fully specified by

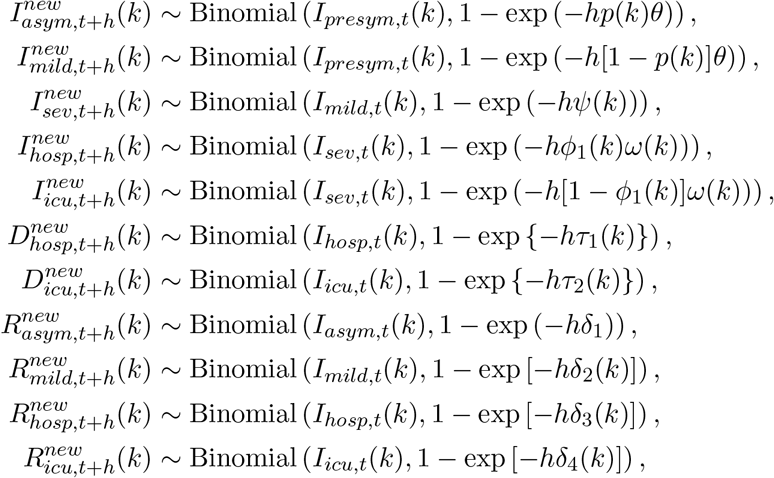

and

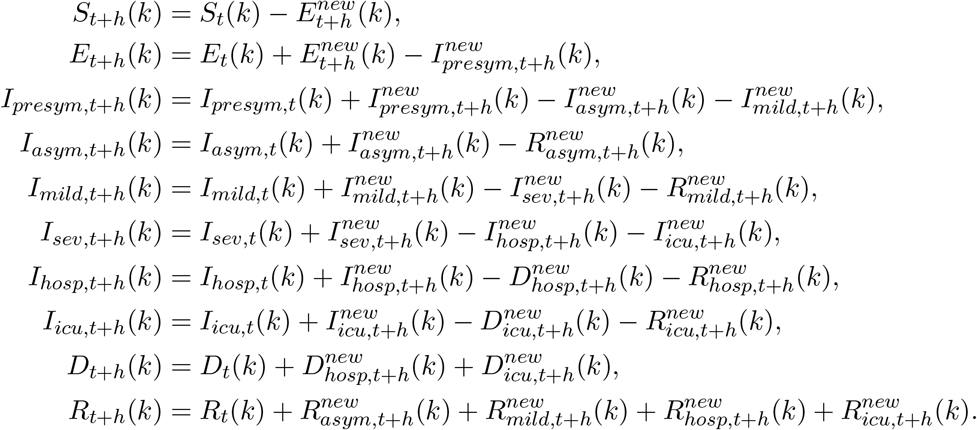

Predictions based on the stochastic discrete age-structured epidemic model will account for two sources of variability, namely (1) variability coming from the observational process reflected in uncertainty about the model parameters; and (2) variability introduced by the stochastic process. An overview of the fixed parameter values, sources (incl. literature), and distributional assumptions are listed in Table B3 of Appendix B.

### 2.4 Next generation matrix and basic reproduction number

The basic reproduction number *R*_0_ for the proposed compartmental model can be obtained by means of the next-generation approach [11]. More specifically, the basic reproduction number is equal to the leading eigenvalue of the next generation matrix, i.e., *R*_0_ is

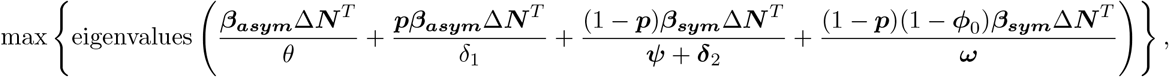

where ***M*** Δ***V*** operates by multiplying the *i*th row of matrix ***M*** with the *i*th element of column vector ***V***. Note that the vector ***N*** ≈ ***S***(0) denotes the population age distribution (i.e., the number of individuals in each age group in the population). The time-dependent effective reproduction *R*_*t*_ is obtained by replacing ***N*** with the number of susceptible individuals ***S***(*t*) at time *t*.

### 2.5 Intervention measures

Intervention measures mainly targeted the reduction of face-to-face contacts as an effective way of breaking the transmission chains of COVID-19 disease. These measures have led to significant alterations in social mixing patterns, hence, changing the trajectory of the dynamics of COVID-19. To assess the impact of these measures, we utilize social contact matrices derived using the online SOCRATES tool [4], developed for social contact data sharing and assessment of mitigation strategies, and including survey data for various locations, i.e, home, work, school, transport, leisure and other places. The imposed measures have changed the contacts made on these respective contact locations and altered disease transmission. Different social contact matrices are considered to describe the data prior to the lockdown measures and those quantifying contact patterns after the interventions taken. Different choices with regard to the reduction in social contacts are as outlined in Table 1 and Appendix C, and their performance in terms of model fit is compared using the Deviance Information Criterion (see Appendix C for specific details). Note that the choice of the intervention matrix quantifies the extent of social contact reductions, thereby determining the reduction in the effective reproduction number following the installment of stringent lockdown measures [12].

**Table 1:** Different social contact matrices considered to quantify the impact of the intervention measures on social contact patterns. Percentage of average number of pre-pandemic contacts at different locations. WT: Work and transport reductions, SC: School closure.

| Social contact matrix | Work & Transport | School closure | Leisure & other activities |
| --- | --- | --- | --- |
| 50% WT & SC | 50% | Yes | 10% |
| 60% WT & SC | 40% | Yes | 10% |
| 70% WT & SC | 30% | Yes | 10% |
| 80% WT & SC | 20% | Yes | 10% |
| 90% WT & SC | 10% | Yes | 10% |

Compliance to the intervention measures taken is assumed to be gradual and is therefore modelled in a flexible way. More specifically, we consider a logistic compliance function

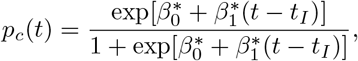

where *t*_*I*_ is the time at which the interventions are initiated. The slope parameter associated with the compliance function (i.e., 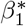) is estimated based on the available data.

### 2.6 Exit strategies

Following the intervention measures that the Belgian government imposed towards limiting the spread of COVID-19 disease, well-tailored exit strategies are needed in order to enable individuals individuals to resume their normal social life whilst protecting the health care system from unprecedented pressure leading to unnecessary loss of lives. Here, we explore and compare possible approaches in lifting imposed measures. The different aspects within the exit strategies are listed below:

- Progressive lifting of lockdown measures on key sectoral pillars of the economy that require physical presence for workers/staff while keeping non-essential service providers closed. This will entail progressively re-adjusting the social contact matrices made at work, during travel/transport, and contacts at other places.
- Gradual re-opening of schools. This will entail re-adjusting the contacts made at school. In line with the exit strategies adopted in Belgium, various partial re-opening dates are considered and their joint impact is explored.
- Opening social places like restaurants, retail stores and hotels. This will involve re-adjusting the social contact matrices for those contacts made during leisure activities, work and transportation as well as those made at other places.

Note that the aforementioned exit strategies cannot be looked at independently since, e.g., parents going back to work will have to rely on childcare/schools to take care of their children. Therefore, we will refer to exit scenarios rather than individual strategies in the remainder of the paper.

To assess the effectiveness of the individual exit strategies and combined scenarios, several comparisons will be made as follows: each exit scenario will be compared to a (baseline) situation without changes, and with each other. More details on the exit scenarios and the translation to the relative number of contacts compared to the pre-pandemic situation are outlined in Table 2. The different scenarios presented in this table give rise to a gradual relief of the lockdown measures taken, similar to the current strategy in Belgium:

**Table 2:** Exit scenarios considered in combination with the best fitting intervention social contact matrix without any lifting of the measures taken as the baseline scenario. Differences in contact percentages in subsequent phases highlighted in bold for each scenario.

| Scenario | Timing | Work & Transport | School | Leisure & other activities |
| --- | --- | --- | --- | --- |
| Baseline | – | 20% | 0% | 10% |
| S1 | Phase 1a-1b | **30%** | 0% | **20%** |
| S2 | Phase 1a | **30%** | 0% | **20%** |
|  | Phase 1b | **40%** | 0% | **30%** |
| S3 | Phase 1a | **30%** | 0% | **20%** |
|  | Phase 1b | **50%** | 0% | **40%** |
| S4 | Phase 1a-1b | **40%** | 0% | **30%** |
|  | Phase 2a-2b | 40% | **20%** | 30% |
| S5 | Phase 1a-1b | **40%** | 0% | **30%** |
|  | Phase 2a-2b | 40% | **40%** | 30% |
| S6 | Phase 1a-1b | **40%** | 0% | **30%** |
|  | Phase 2a-2b | 40% | **60%** | 30% |
| S7 | Phase 1a-1b | **40%** | 0% | **30%** |
|  | Phase 2a-2b | 40% | **20%** | 30% |
| S8 | Phase 1a-1b | **40%** | 0% | **30%** |
|  | Phase 2a | 40% | **20%** | 30% |
|  | Phase 2b | 40% | **40%** | 30% |
| S9 | Phase 1a-1b | **40%** | 0% | **30%** |
|  | Phase 2a | 40% | **20%** | 30% |
|  | Phase 2b | 40% | **60%** | 30% |
| S10 | Phase 1a-1b | **40%** | 0% | **30%** |
|  | Phase 2a-2b | 40% | **20%** | 30% |
|  | Phase 3 | 40% | 20% | **20%** |
| S11 | Phase 1a-1b | **40%** | 0% | **30%** |
|  | Phase 2a-2b | 40% | **20%** | 30% |
|  | Phase 3 | 40% | 20% | 30% |
| S12 | Phase 1a-1b | **40%** | 0% | **30%** |
|  | Phase 2a-2b | 40% | **20%** | 30% |
|  | Phase 3 | 40% | 20% | **40%** |

Phase 1a – May 4: Although remote work remains the norm, business-to-business services and companies that are able to comply with physical distancing measures re-opened;

Phase 1b – May 11: Shops re-opened under strict requirements related to organisation of the work and restricting access to the store to avoid overcrowding;

Phase 2a – May 18: Schools partially re-opened (first phase - selected grades in primary and secondary schools);

Phase 2b – June 2: Schools partially re-opened further (second phase - pre-primary schools);

Phase 3 – June 8: Restaurants, bars, and cafes re-opened under strict measures including physical distancing and a limited number of customers;

These comparisons were mainly made on the basis of the daily number of new hospitalizations and admissions to the ICU. Furthermore, the implementations thereof in combination with the timing of holiday and school periods will be studied to explore whether rebound effects will occur, i.e., whether deconfinement results in subsequent COVID-19 waves.

### 2.7 Data and estimation procedure

In this section we briefly describe the parameter estimation procedure and different data sources that are considered to fit the models.

#### 2.7.1 Deterministic model

We first fit the deterministic version of the proposed compartmental model to the initial phase of the epidemic. More specifically, we fit the model to the daily numbers of new COVID-19 hospitalizations (for all age groups combined) starting from 1 March 2020 until 22 March 2020 (before intervention measures had an influence on hospital admissions). We use a likelihood-based approach by assuming

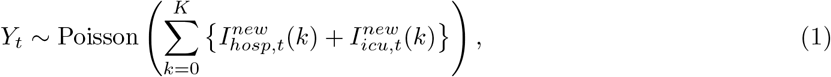

where the realization *y*_*t*_ of *Y*_*t*_ is the observed total number of new hospitalizations across all age groups at time (day) *t* (i.e., within the last 24 hours), and 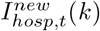 and 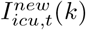 are the expected number of new hospitalizations (without ICU) and ICU admissions at time *t* in age group *k*, respectively. These expected numbers are obtained by numerically solving the set of ordinary differential equations. The aforementioned procedure is used to obtain reasonable starting values for the model parameters in order to initialize the MCMC sampler (see Section 2.7.4 in fitting the stochastic compartmental model to the available outbreak data).

#### 2.7.2 Stochastic model

Next to the social contact data used to inform the transmission parameters, three different data sources are considered when fitting the stochastic compartmental model, namely (1) age-specific data on the daily number of new hospitalizations (until May 4) [13]; (2) age-specific data on the daily number of new deaths (excluding deaths in elderly homes) (until May 4) [13]; and (3) serial serological survey data collected during the epidemic [5]. Belgian hospitals are obliged to report the daily number of new hospitalizations to the Scientific Institute of Public Health, Belgium (Sciensano), which are made publicly available through an online platform [13]. Age-specific hospitalization data were collected through the clinical surveillance database of COVID-19 hospitalized patients [14]. This database is an ongoing multicenter registry collecting information on hospital admission related to COVID-19 infection. Patient-specific characteristics are collected through two online questionnaires: one related to admission and one related to discharge. As the reporting is strongly recommended by the Belgian Risk Management Group, the reporting coverage is high including more than 70% of all hospitalized COVID-19 cases during the first wave [14]. Based on this information, the weekly age distribution of hospitalized cases (see Figure D1 in Appendix D) is derived such that the total daily incidence of hospitalizations is transformed to be age-specific. Reporting of the daily incidence of COVID-19 related deaths by age within hospitals is mandatory and made publicly available on the Sciensano dashboard [13]. The (serial) serological data is obtained from two data collections (30 March – 5 April, 2020 & 20 April – 26 April, 2020) within a prospective cross-sectional seroprevalence study and based on residual sera obtained from individuals aged 0-101 years. Seropositivity of the samples is determined based on a semi-quantitative ELISA test kit (EuroImmun, Luebeck, Germany) measuring IgG antibody concentrations against S1 proteins of SARS-CoV-2 in serum (see Appendix E for more details).

The following distributional assumptions are made with regard to the different outcome variables:

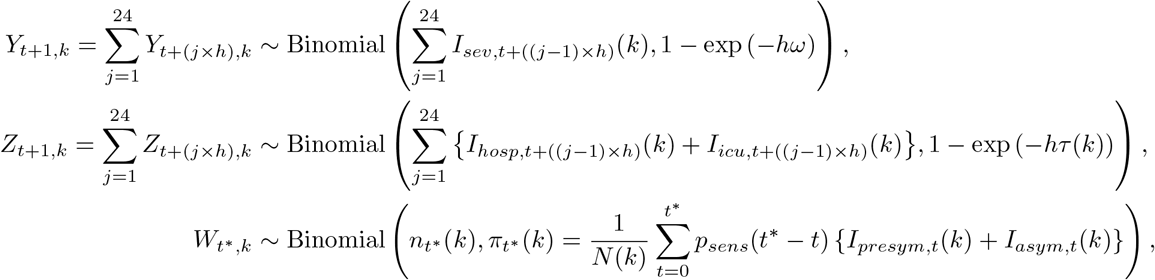

where *Y*_*t,k*_ and *Z*_*t,k*_ represent the number of new hospitalizations and new deaths at time *t* in age group *k*, respectively, relying on the assumption of equal age-specific mortality rates *τ*_1_(*k*) = *τ*_2_(*k*) *≡ τ* (*k*) for hospitalized patients on general and ICU wards. Since we do not have data on referral within hospitals, we do not explicitly distinguish between hospitalized and ICU admitted patients in terms of hospital discharge (including death), although the model is equipped to do so.

Moreover 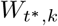 represents the total number of seropositive individuals in age group *k* in a cross-sectional serological collection of residual blood samples performed at time *t*^*^. All individuals tested in age group *k* at time *t*^*^, denoted by 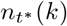, have a probability 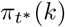 (i.e., equal to the observed seroprevalence) to be classified as seropositive accounting for sensitivity of the test *p*_*sens*_(*t*_*o*_) as a function of time since symptom onset and assuming perfect specificity of the test. The sensitivity of the test is assumed to follow a logistic growth curve based on available information in the literature [15]. For more details, the reader is referred to Appendix E. Weighted seroprevalence estimates are used in the analysis [5].

#### 2.7.3 Model initialization

The number of imported cases (and first generation(s) of infected cases through local transmission) is determined from the age-specific number of confirmed cases on 12 March 2020. More specifically, given a number *n*_0_(*k*) of confirmed cases in age group *k*, the expected number of imported cases in age class *k* equals

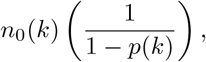

where *p*(*k*) represents the asymptomatic fraction in age group *k* thereby assuming that confirmed cases solely reflect the proportion of mildly and severely ill individuals. The introduction of the imported cases in the system is presumed to take place on 1 March following the school holiday period.

#### 2.7.4 Estimation

Model parameters are estimated using a Markov Chain Monte Carlo (MCMC) approach. A two phase method is considered in which the first phase consists of an adaptive Metropolis-within-Gibbs (AMWG) [16, 17] and/or adaptive mixture Metropolis-Hastings (AMM) algorithm [17] to achieve stationary samples that seem to have converged to the target posterior distributions (stationarity is obtained after a maximum of 250,000 iterations). In the second phase, a non-adaptive Random-Walk Metropolis (RWM) algorithm [18] is used to draw final samples from the posterior distributions. More specifically, 500,000 iterations were conducted thereby retaining every 100th iteration after discarding an initial burn-in part of 250,000 iterations. An overview of the different prior distributions is presented in Table B4 of Appendix B. In order to ensure that plausible parameter values are obtained, logit- and log-transformations are considered depending on the required range for the different model parameters.

## 3 Results

In this section, we show the results of fitting the stochastic compartmental model to the data at hand. First of all, we study the fit to the data and the posterior distributions of the model parameters. Next, we investigate the age- and time-varying (sero)prevalence derived from the model. Finally, we investigate the impact of different exit strategies on the resurgence of the COVID-19 epidemic.

### 3.1 Baseline scenario accounting for mitigation strategies

Both the probability of experiencing an asymptomatic infection and the probability of having mild symptoms upon contracting COVID-19, i.e., ***p*** and ***ϕ***_0_ are assumed to be age-dependent. The latter is estimated using a prior distribution based on current literature (see Tables B3 and B4 for more details). The relative infectiousness of asymptomatic versus symptomatic individuals *r*_*β*_ is fixed at a value of 0.51 [19]. Other model parameters are either fixed or estimated based on the available data (see Table B3 and Appendix F).

The best fitting model included social contact matrices with an 80% reduction of the normal work and transportation contacts (*α* = 0.2 in Appendix C), with no school contacts and with 10% of the regular leisure contacts and contacts related to other activities. In Figure 2, we graphically depict 25 stochastic realizations of the hospital admissions and deaths since March 1 based on a thinned chain from the joint posterior distribution of the model parameters together with pointwise 95% credible intervals derived from stochastic realizations based on 5000 random draws from the joint posterior distribution of the model parameters. The figure clearly shows that the observed daily number of hospitalizations and deaths (black dots) are well described by the model. Furthermore, the estimated age-dependent daily numbers of new hospitalizations and deaths are graphically depicted in Figure 3 for the 10 age categories.

**Figure 2:**
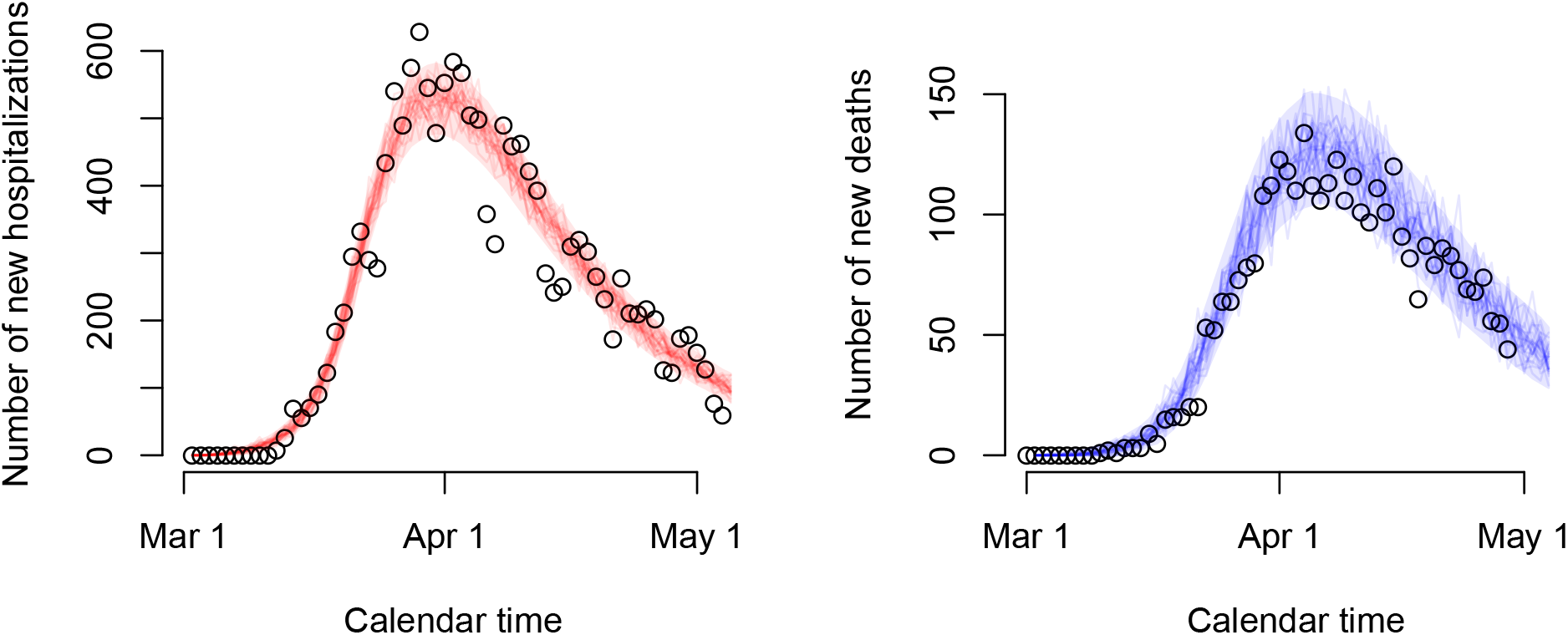
Stochastic realizations of the compartmental model based on a thinned MCMC chain from the joint posterior distribution of the model parameters and relying on an ‘asymptomatic’ and ‘symptomatic’ social contact matrix composed of 20% of regular work and transportation contacts, no school contacts and 10% of leisure contacts and contacts related to other activities. Shaded areas represent 95% credible intervals. Reported daily number of hospitalizations and deaths are represented by black circles.

**Figure 3:**
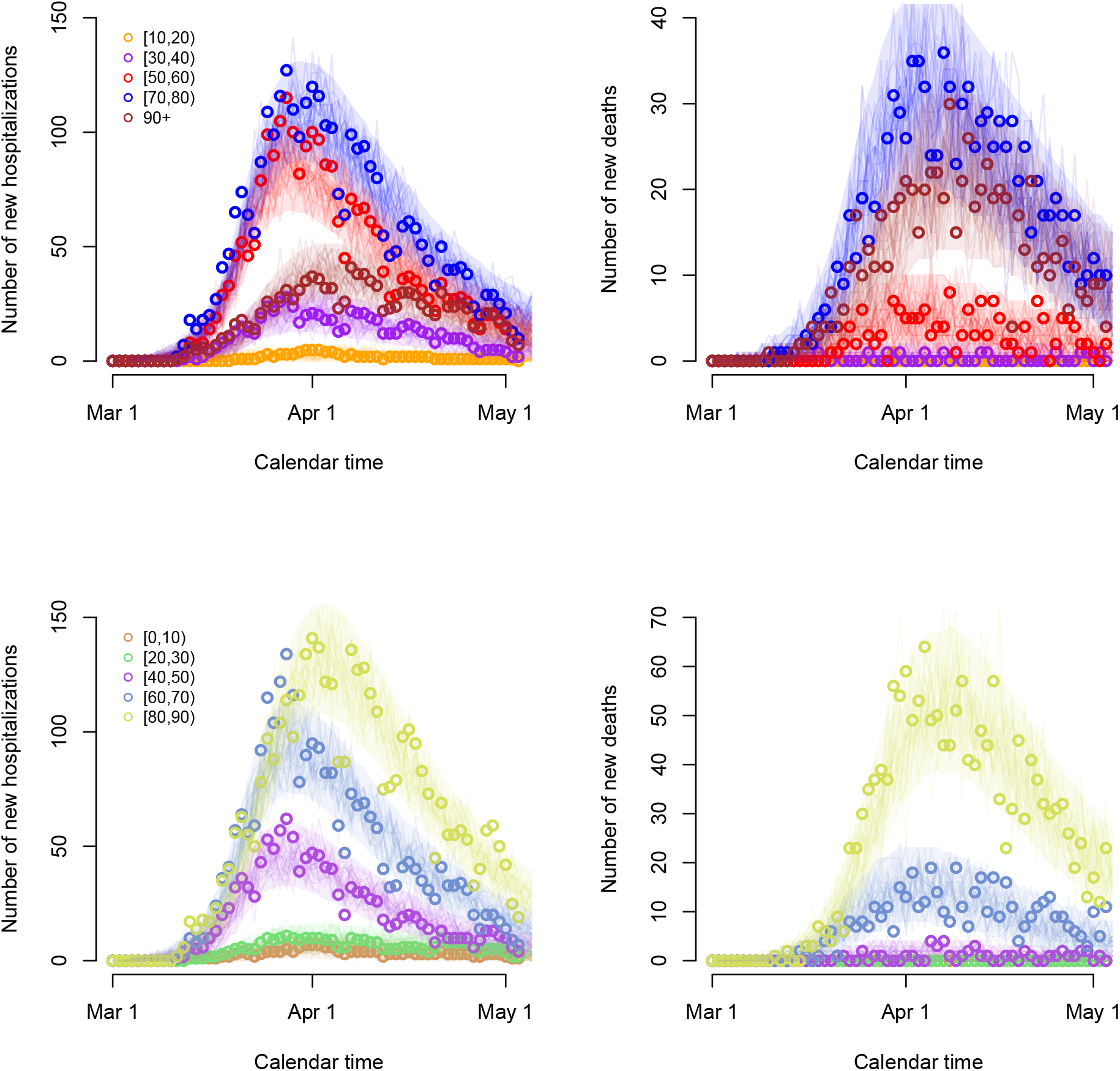
Stochastic realizations of the compartmental model based on a thinned MCMC chain from the joint posterior distribution of the model parameters and relying on an ‘asymptomatic’ and ‘symptomatic’ social contact matrix composed of 20% of regular work and transportation contacts, no school contacts and 10% of leisure contacts and contacts related to other activities. Number of new hospitalizations (left upper and lower panels) and deaths (right upper and lower panels) are shown for all 10 age groups. Shaded areas represent 95% credible intervals. Reported daily number of hospitalizations and deaths are represented by circles.

### 3.2 Posterior distributions

In Table 3, we present summary measures for the posterior distributions of the most important (implicit) model parameters including the posterior mean, median, standard deviation and 95% credible intervals (CIs). An overview of posterior quantities for all model parameters is included in Appendix F).

**Table 3:**
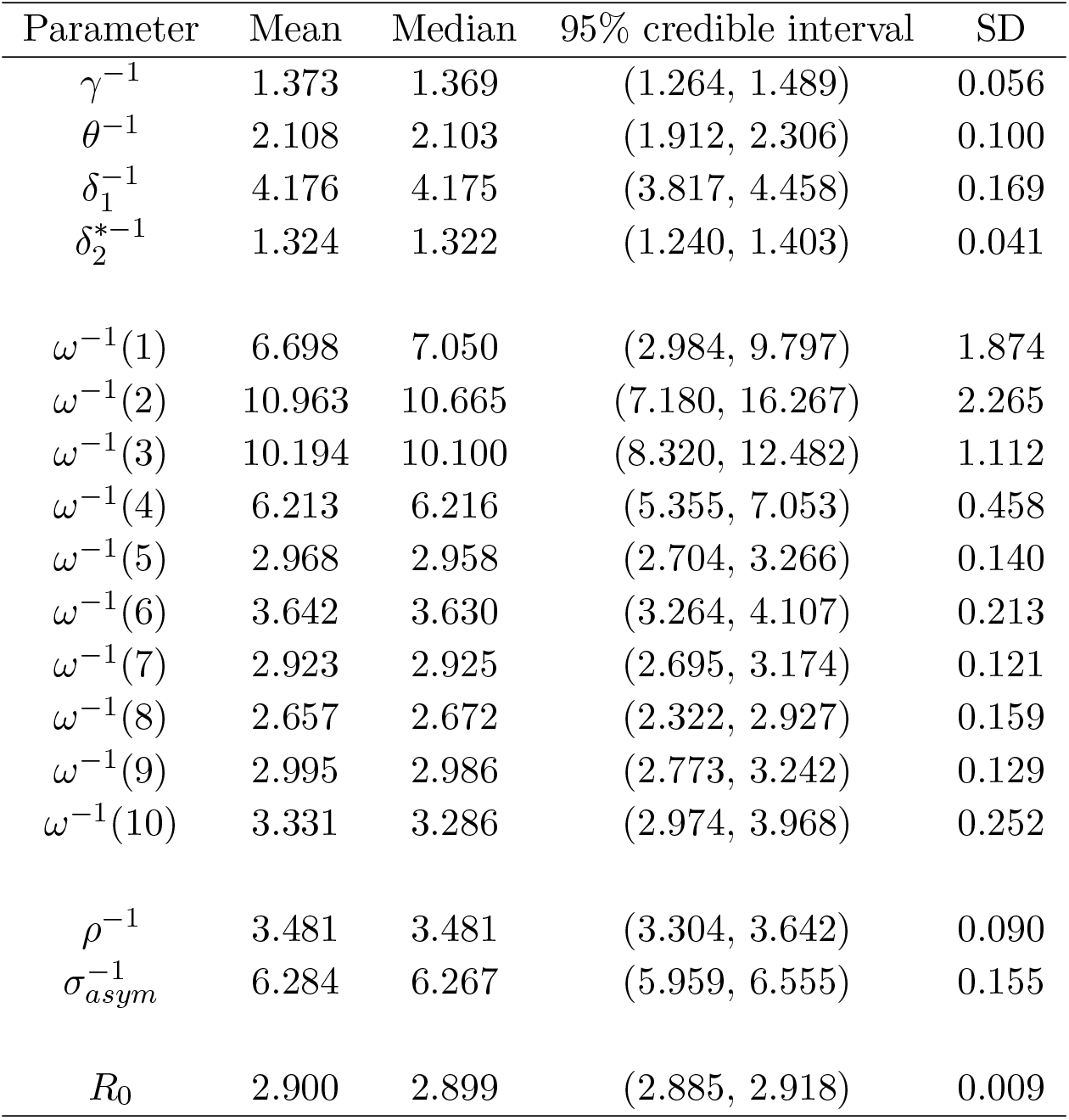
Posterior mean, median, standard deviation (SD) and 95% credible interval for the model parameters.

| Parameter | Mean | Median | 95% credible interval | SD |
| --- | --- | --- | --- | --- |
| $\gamma^{-1}$ | 1.373 | 1.369 | (1.264, 1.489) | 0.056 |
| $\theta^{-1}$ | 2.108 | 2.103 | (1.912, 2.306) | 0.100 |
| $\delta_1^{-1}$ | 4.176 | 4.175 | (3.817, 4.458) | 0.169 |
| $\delta_2^{*-1}$ | 1.324 | 1.322 | (1.240, 1.403) | 0.041 |
| $\omega^{-1}(1)$ | 6.698 | 7.050 | (2.984, 9.797) | 1.874 |
| $\omega^{-1}(2)$ | 10.963 | 10.665 | (7.180, 16.267) | 2.265 |
| $\omega^{-1}(3)$ | 10.194 | 10.100 | (8.320, 12.482) | 1.112 |
| $\omega^{-1}(4)$ | 6.213 | 6.216 | (5.355, 7.053) | 0.458 |
| $\omega^{-1}(5)$ | 2.968 | 2.958 | (2.704, 3.266) | 0.140 |
| $\omega^{-1}(6)$ | 3.642 | 3.630 | (3.264, 4.107) | 0.213 |
| $\omega^{-1}(7)$ | 2.923 | 2.925 | (2.695, 3.174) | 0.121 |
| $\omega^{-1}(8)$ | 2.657 | 2.672 | (2.322, 2.927) | 0.159 |
| $\omega^{-1}(9)$ | 2.995 | 2.986 | (2.773, 3.242) | 0.129 |
| $\omega^{-1}(10)$ | 3.331 | 3.286 | (2.974, 3.968) | 0.252 |
| $\rho^{-1}$ | 3.481 | 3.481 | (3.304, 3.642) | 0.090 |
| $\sigma_{asym}^{-1}$ | 6.284 | 6.267 | (5.959, 6.555) | 0.155 |
| $R_0$ | 2.900 | 2.899 | (2.885, 2.918) | 0.009 |

The basic reproduction number *R*_0_ at the start of the epidemic - prior to any government intervention - is estimated to be 2.900 with 95% CI (2.885, 2.918). On May 4, 2020 the effective reproduction number after the lockdown was estimated to be 0.738 (95% CI: 0.732, 0.744). The time-dependent effective reproduction number *R*_*t*_ is shown in Figure F3 of Appendix F. The posterior means for the parameters *ω*^(^ − 1)(*k*) are in line with estimates of the median duration between symptom onset and hospitalization [20]. Furthermore, the average length of the incubation and asymptomatic infectious period, *ρ*^−1^ and 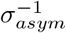 are estimated to be 3.481 days (95% CI: 3.304, 3.642) and 6.284 days (95% CI: 5.959, 6.555), respectively, very similar to values reported in the literature (see Table B3). For individuals experiencing severe symptoms, the average length of the infectious period, constrained due to isolation in hospital, depends on the age-specific time between symptom onset and hospitalization. Full compliance to the intervention measures was obtained after approximately 6 days (see Appendix F).

Boxplots of the marginal posterior distributions of the probability of hospitalization are presented in Figure 4. The probability of hospitalization increases substantially with increasing age. A decrease in hospitalization probability is observed in age class [80, 90) after which it increases again for individuals of age 90+. However, the probability of hospitalization, as a proxy of disease severity, is likely time-dependent as well as biased for the oldest age groups due to differential referral policy in elderly homes or end of life choices in the last will of severely ill persons. More specifically, the general population essentially consists of two subpopulations, i.e., a nursing home and non-nursing home population. It is likely that in reality, the nursing home residents were more often exposed than elderly in the general population of the same age. One other explanation for the decrease in probability of hospitalization is that nursing home residents (which constitute an important part of the age group between 80 and 89 years of age) were less likely referred to the hospital (as would have been the case when they were not living in a nursing home), at least during the initial part of the epidemic, thereby implying a lower hospitalization probability in that age group. Moreover, persons in age class [80, 90) living in the non-nursing home population are believed to be in better condition than nursing home residents of that age, being more frail when suffering from more severe comorbidities. Hence, this could lower the probability of hospitalization further. Estimated mortality and infection fatality rates are shown in Figure F4 in Appendix F.

**Figure 4:**
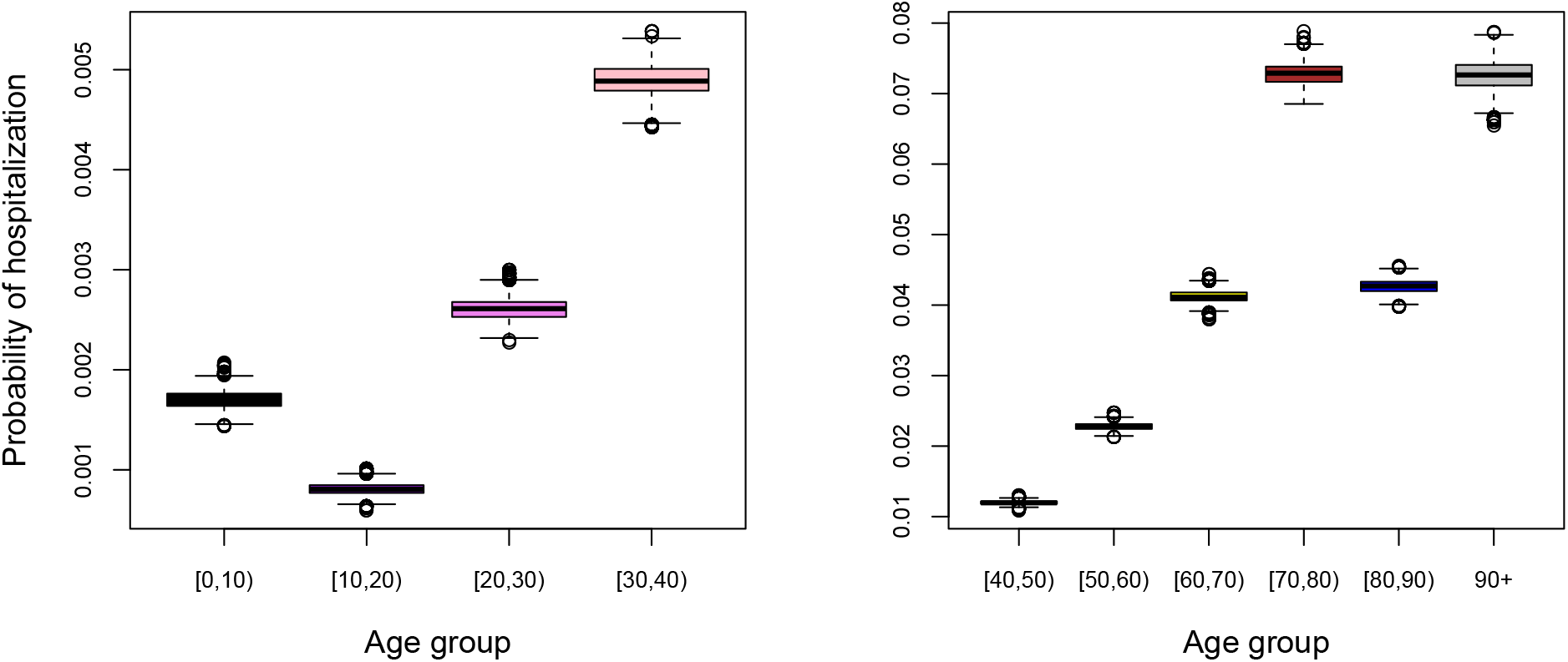
Boxplots of the marginal posterior distributions of the probability of hospitalization by age group.

### 3.3 Estimated age- and time-dependent (sero)prevalence of COVID-19

In Figure 5, we show the estimated and observed seroprevalence on March 30, 2020 (left panel) and on April 20, 2020 (right panel) with 95% credible intervals in gray dashed lines and asymptotic 95% error bars for the weighted seroprevalence.

**Figure 5:**
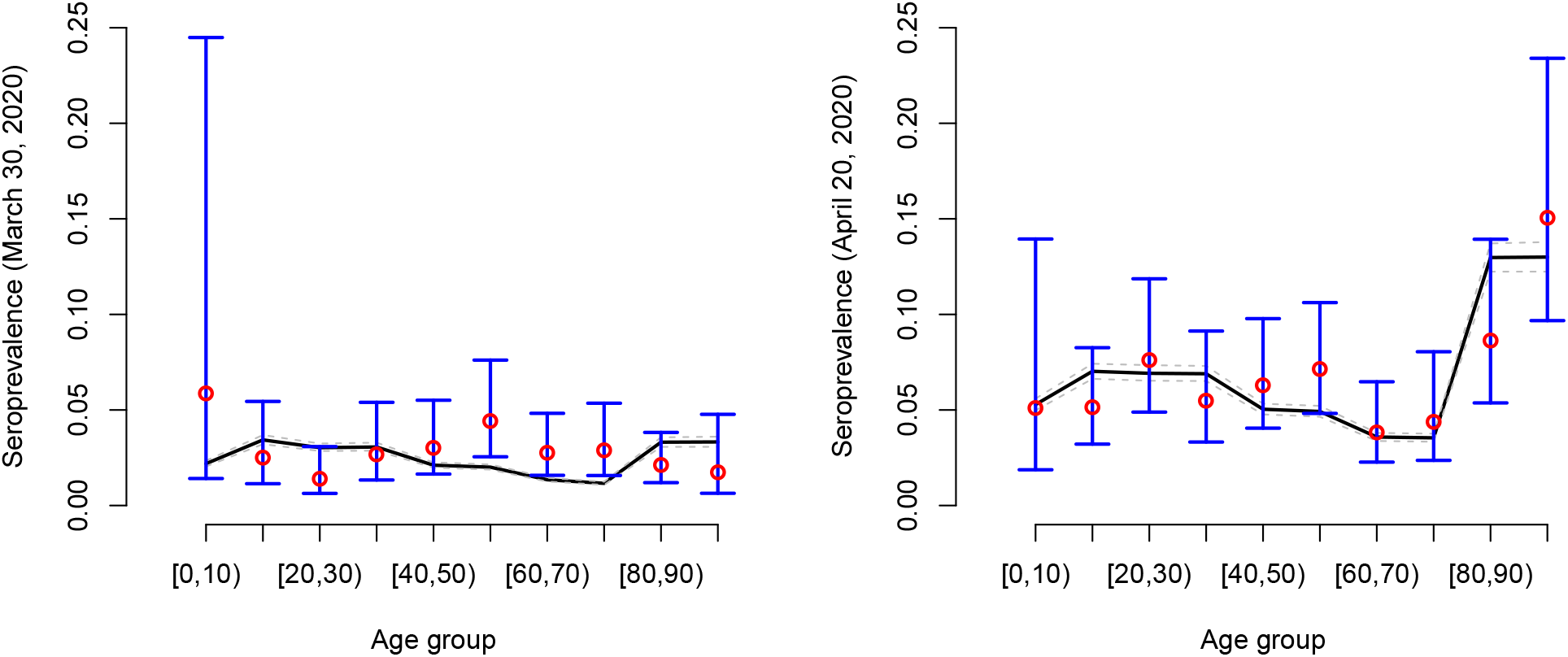
Estimated age-dependent seroprevalence of COVID-19 with 95% credible interval on March 30, 2020 (left panel) and April 20, 2020 (right panel). Observed seroprevalences are depicted using red dots with 95% confidence intervals in blue. The confidence interval for the age group [0, 10) is wide due to the low number of individuals (*n* = 36).

The estimated age- and time-dependent prevalence of COVID-19 in the Belgian population is shown in Figure 6. Clearly, the estimated prevalence is higher in the oldest age groups ([80,90), 90+) which is in line with the observed seroprevalence, the latter providing a cross-sectional snapshot of the (delayed) build-up of seropositivity in the population upon infection (see Figure 5). The estimated overall weighted prevalence of COVID-19 is equal to 0.069 (95% CI: 0.064, 0.073) on May 4, 2020 (black dashed line with gray shaded area).

**Figure 6:**
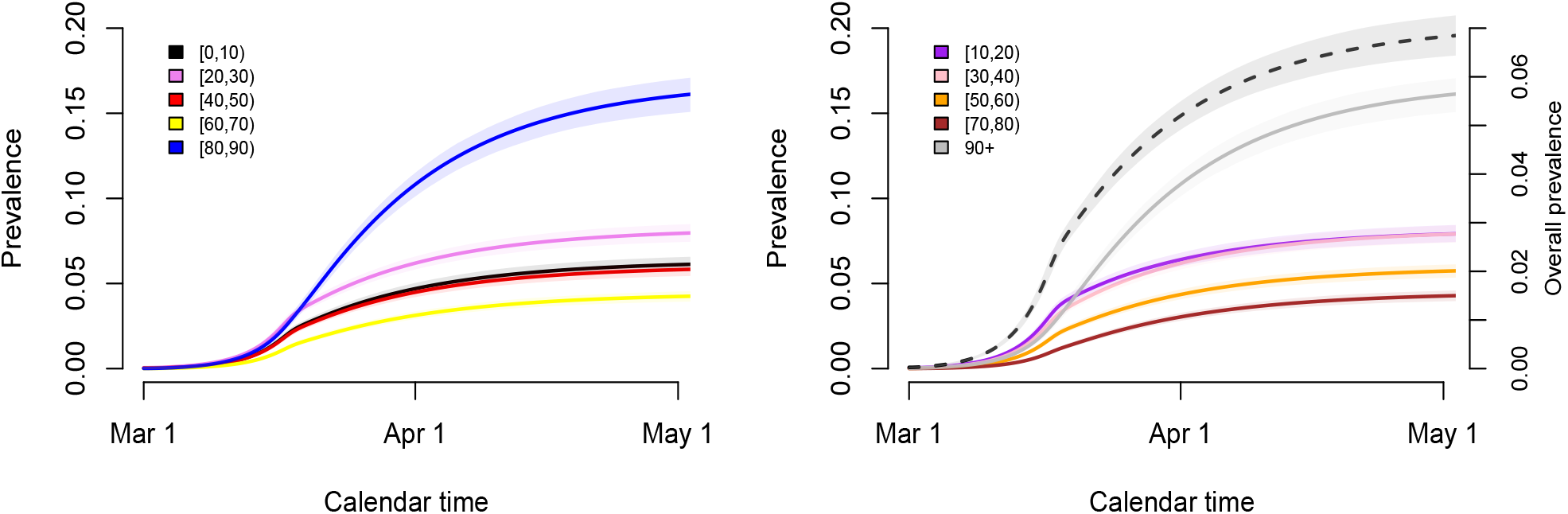
Estimated time-dependent prevalence of COVID-19 in the different age groups and its weighted average (right panel; black dashed line - right y-axis) based on 5000 stochastic realizations given random draws from the joint posterior distribution of the model parameters with 95% credible intervals (shaded regions).

### 3.4 Exit strategies

We performed short-term predictions of the dynamics of COVID-19 in the Belgian population through the use of scenario analyses. In Figure 7, we show the impact of relaxing lockdown measures by increasing contacts made by people at different locations. More specifically, we present stochastic realizations of the model to predict the number of new hospitalizations under different scenarios with changes in contact behaviour as of May 4, 2020 (Phase 1a, Scenarios S1–S3) and May 11, 2020 (Phase 1b). From May 4 onward, we presume that work-related contacts will increase from 20% (baseline scenario - see Table 2) to 30% of the pre-pandemic number of contacts at work (or transmissibility is reduced to an extent equivalent with the assumed reduction in work-related contacts), and that the number of transport contacts and contacts during leisure and other activities will increase respectively from 20% to 30% and from 10% to 20% of their pre-pandemic values. On top of that, work, transport and leisure contacts stay constant (scenario S1) or increase to 40%, 40% and 30% (S2) or 50%, 50% and 40% (S3) of the pre-pandemic values, respectively, from May 11 onward. Moreover, a delay of one week is considered for each change in social contact behaviour (i.e., a full extent of all changes in behaviour is reached on May 18, 2020). A small to moderate increase in the contacts at work, transportation and during leisure (blue and purple scenarios) leads either to a complete reduction of hospitalizations or a constant number of new hospitalizations over time. Only the most extreme increases in contacts give rise to a resurgence of COVID-19 implying a second wave of COVID-19 infections (scenario S3 - orange lines).

**Figure 7:**
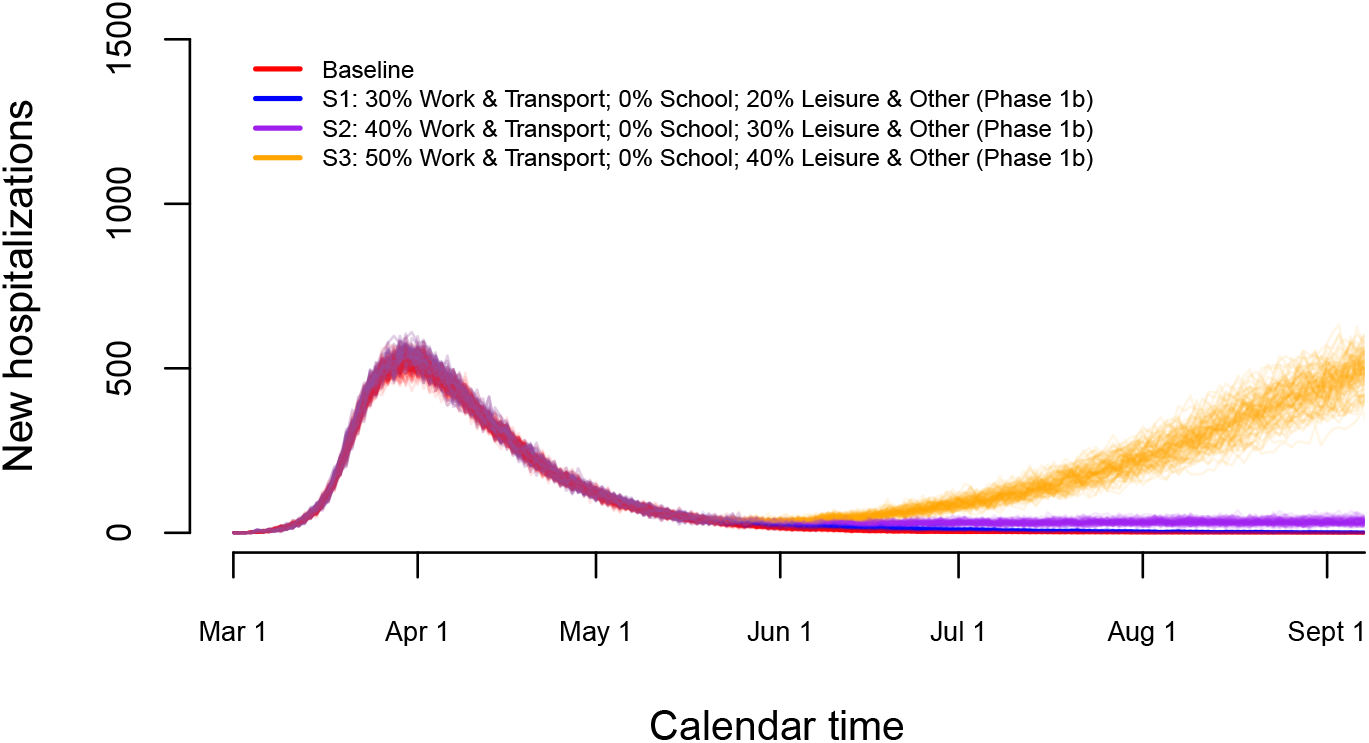
Impact of various exit strategies in terms of the number of work- and leisure-related contacts on the number of new hospitalizations in the absence of re-opening of schools.

A partial re-opening of schools as of May 18 (Phase 2a) is studied in detail in Figure 8. Work- and transport-related contacts and contacts during leisure and other activities increase as of May 4. School-related contacts are assumed to be 20% (S4 - blue lines), 40% (S5 - purple lines) or 60% (S6 - orange lines) of such contacts prior to the epidemic. This increase in school-related contacts is imposed between May 18, 2020 and July 1, 2020. The start of the summer holiday on July 1, 2020 implies a reduction of all school-related contact to 0%. A partial re-opening of schools in combination with a moderate increase in work, transportation and leisure activities leads to a small to moderate increase in the number of new hospitalizations after lockdown measures are relaxed.

**Figure 8:**
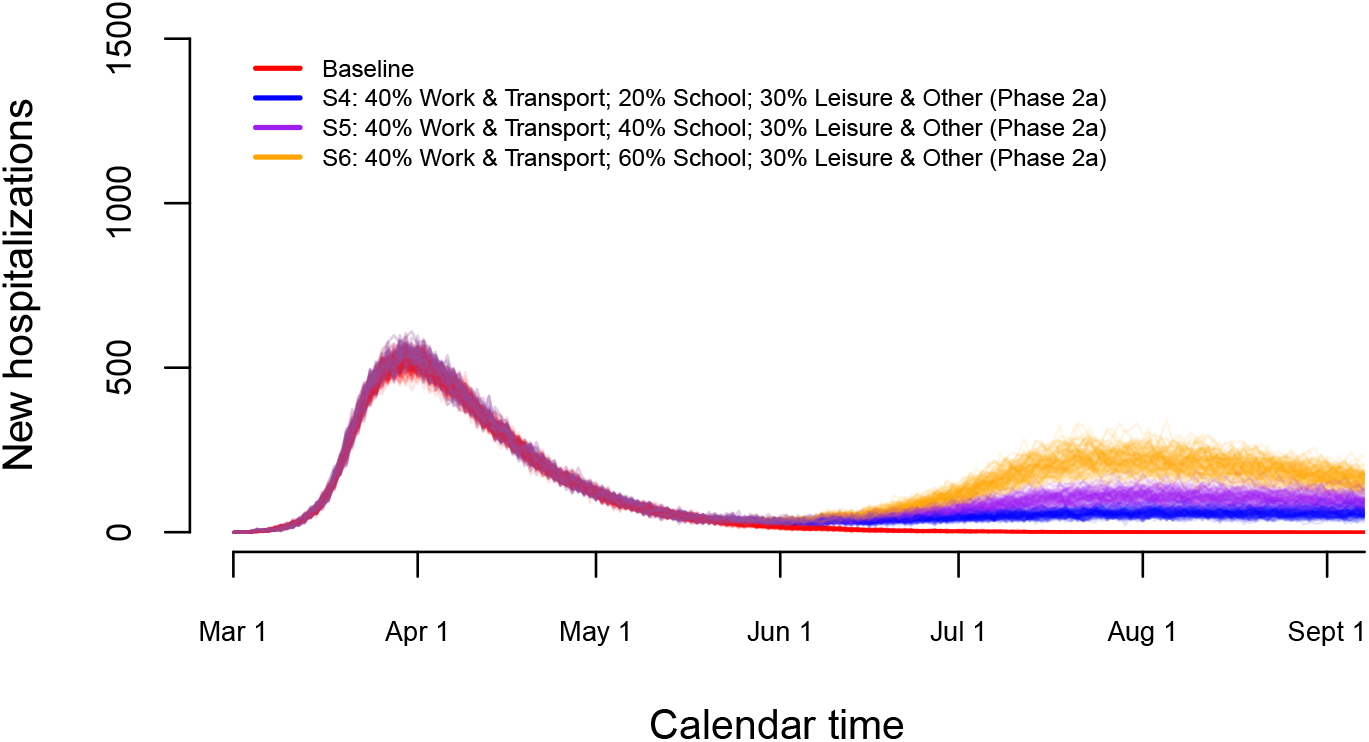
Impact of partial re-opening of schools on the number of new hospitalizations.

Finally, we investigate long-term predictions of subsequent COVID-19 waves for a selection of possible exit scenarios (Figure 9). In those scenarios, we mimic the timing of the Belgian exit strategy. More specifically, schools are partially re-opened on May 18 and June 2, 2020 yielding 20% of school contacts as of May, 18 (S7), an increase from 20% to 40% or 60% of school contacts between May 18 and June 2 for scenarios S8 and S9, respectively. Schools are closed during the vacation period starting from July 1, 2020 until August 31, 2020. We assume that contact behaviour at schools following partial re-opening on September 1, 2020 is equivalent to 60% of the pre-pandemic social contacts made at school. In the lower panel of Figure 9, scenarios S10–S12 show the impact of an increase of leisure contacts to 20%, 40% or 60% of pre-pandemic leisure contacts as of June 8, while assuming school-related contacts to be equal to 20% upon Phases 2a and 2b. Under the assumption of unadapted behaviour given a contact, we observe that due to an insufficient depletion of susceptibles during a second wave of COVID-19 infections (or a phase with a stable daily number of new hospitalizations) a large increase in the number of new hospitalizations will occur by the end of the year with a higher peak size if the one of the second wave (or the plateau level) was lower. The cumulative number of hospitalizations over time is presented in Appendix F. Moreover, leisure contacts are important in determining the peak size of the wave at the end of the year (lower panel of Figure 9). In Figure 10, boxplots of the estimated prevalence over time is shown for scenarios S7–S9 and age groups [0, 10), [30, 40), [60, 70), 90+. The largest increase in prevalence between May 1 (baseline) and December 1, 2020 is observed in the highest age category with an average increase ranging between 36.5% and 38.4% across different scenarios. In all age categories, the increase in prevalence is smallest for scenario S7 and highest for scenario S9.

**Figure 9:**
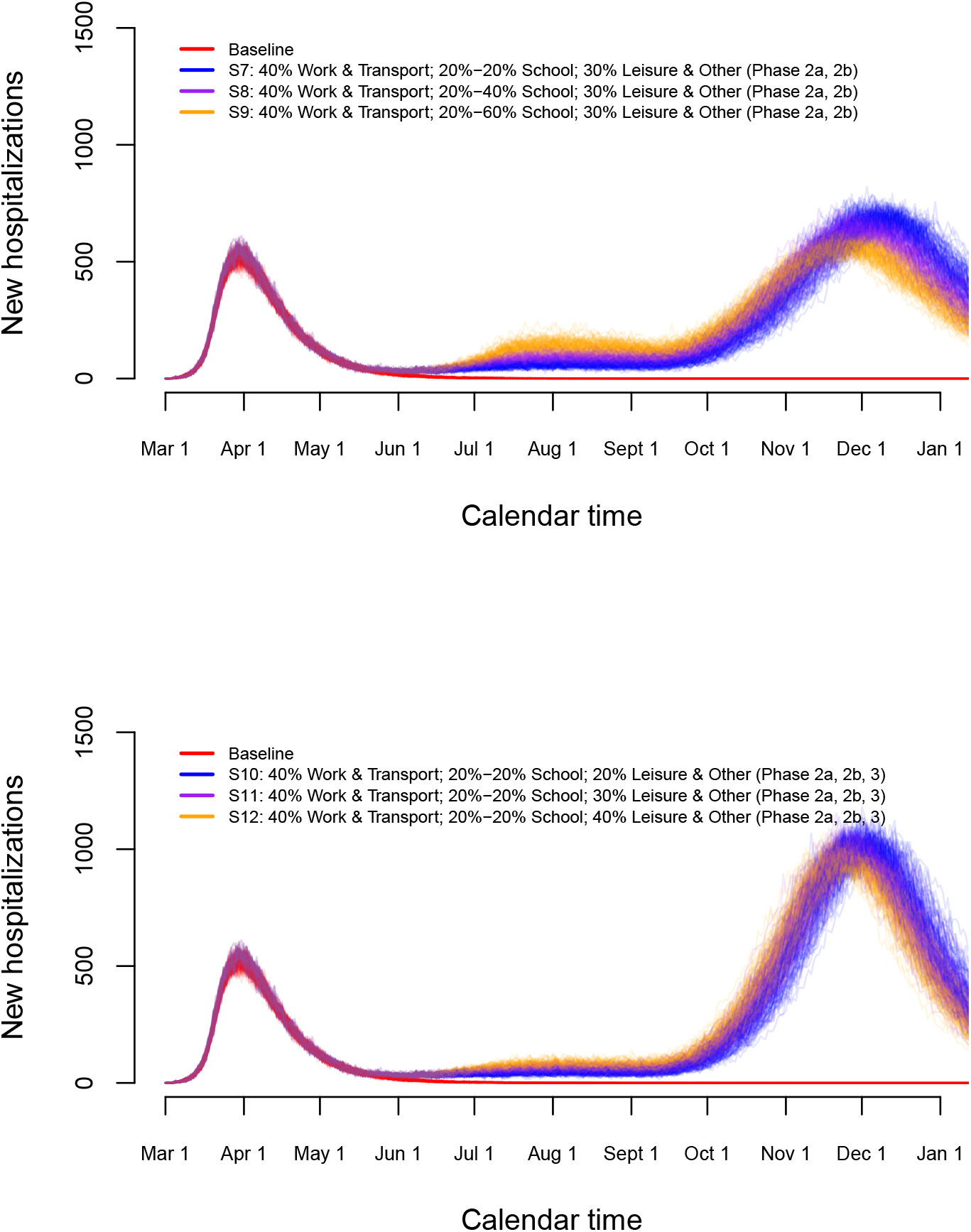
Long-term predictions of the impact of various exit strategies on the number of new hospitalizations.

**Figure 10:**
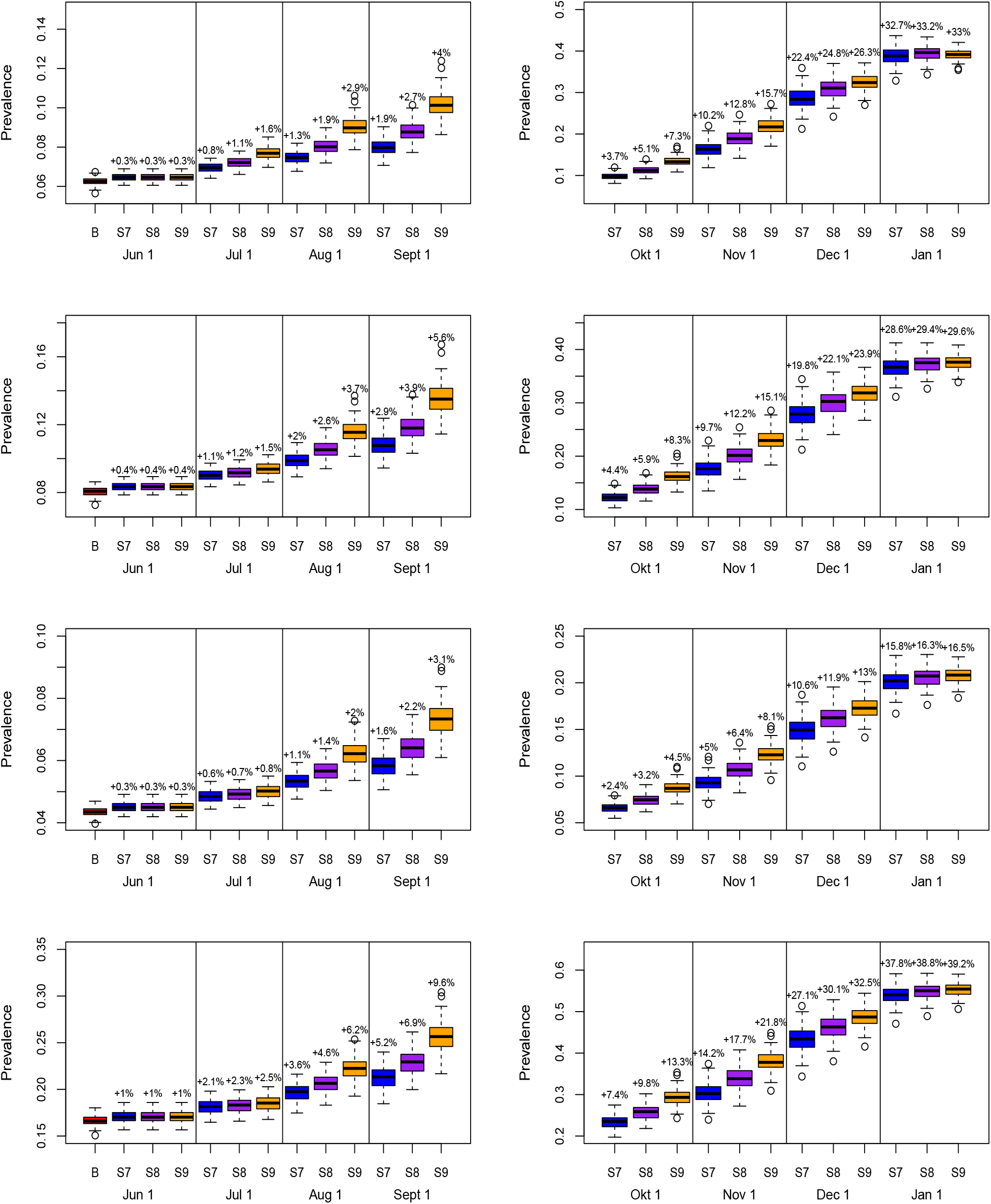
Predictions of the prevalence in exit scenarios S7–S9 for age group [0, 10) (top row), [30, 40) (row 2), [60, 70) (row 3) and 90+ (bottom row). Increments in prevalence compared to the prevalence on May 1, 2020 is added on top of the boxplots.

### 3.5 Validation of the model

Validation of the model is done based on (1) data on new hospitalizations and deaths following the relaxation of the lockdown measures, (2) serological survey data collected in a third round and (3) infection fatality derived from Belgian mortality data [21]. In Figure 11, we show stochastic realizations under the baseline scenario (without change in social contact behaviour after lockdown measures are relaxed) overlaid with new data points after May 4, 2020 (black solid circles). In general, the stochastic model describes the observed data well, even in the absence of changes in contact behaviour after intervention measures were relaxed. Although the observed number of hospitalizations tends to remain constant, thereby deviating from a further decrease thereof in the baseline scenario, no large differences between observed and predicted values are found. Following the gradual relief of the intervention measures, no resurgence of the disease is noticeable to date (end of June).

**Figure 11:**
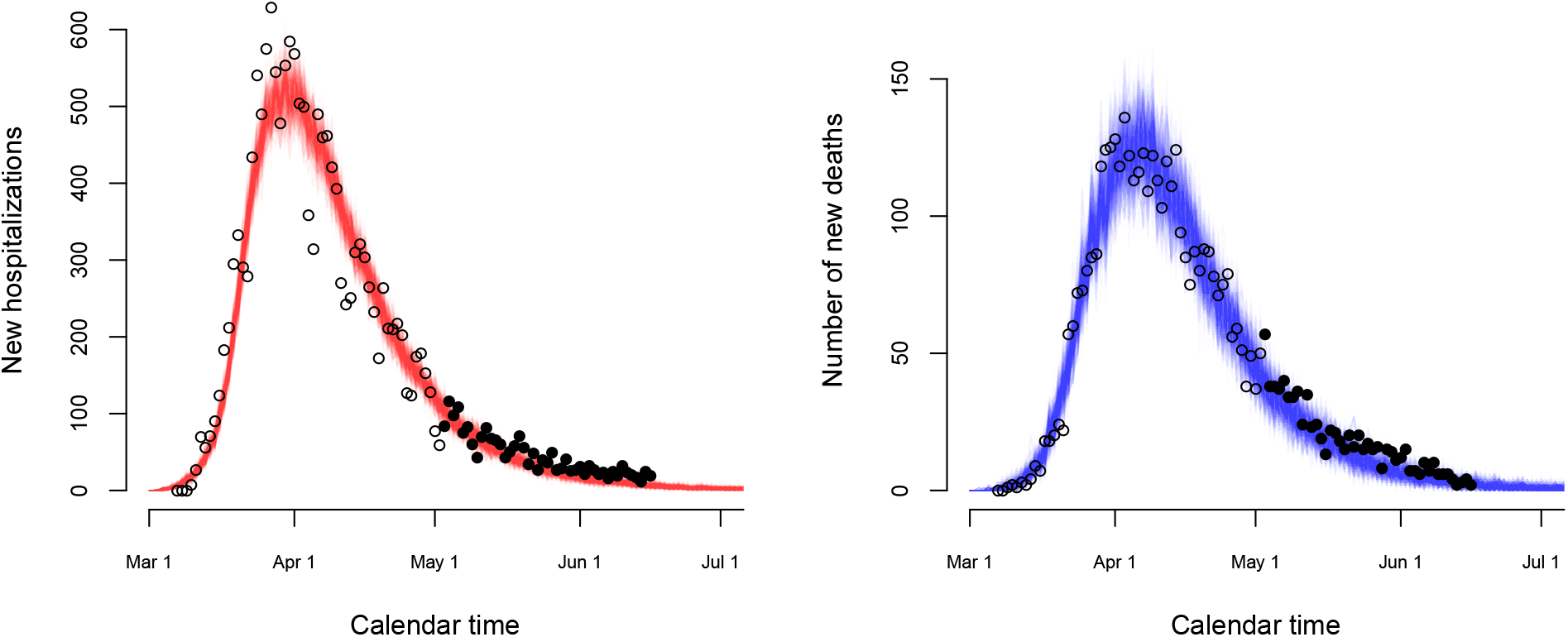
Stochastic realizations based on the baseline scenario (without change in contact behaviour upon relaxing the stringent lockdown measures) for the number of new hospitalizations (left panel) and the number of new deaths (right panel) together with observed data points used for fitting (black open circles) and for validation (black solid circles).

Next, the estimated seroprevalence based on the model is related to the one obtained from a third cross-sectional serological survey. Data was collected in a third round, after the initiation of the gradual relaxation of the stringent measures, between May 18, 2020 and May 25, 2020. The overall weighted seroprevalence was estimated to be 6.87% (95% confidence interval: 5.89%, 8.01%) [5]. In our model, the posterior mean of the seroprevalence is 6.8% with 95% credible interval (6.4%, 7.2%) which is similar to the aforementioned values. Furthermore, the estimated infection fatality rates (IFRs) (see Appendix F) are in line with those reported by Molenberghs et al. (2020) [21]. These authors report an overall IFR of 0.43% (95% confidence interval: 0.30%, 0.62%) in the non-nursing home population whereas our model suggests a posterior mean of 0.507% (95% CI: 0.480%, 0.536%) which is nicely in line. Age-specific IFRs are presented in Appendix F.

## 4 Discussion

In this manuscript, we used a stochastic age-structured discrete time epidemic transmission model fitted to daily hospital admission, COVID-19 related mortality data and serial serological survey data with regard to SARS-CoV-2 antibody presence to describe and study COVID-19 disease dynamics in the Belgian population. As age-specific heterogeneity has been proven to be of great importance in terms of transmission, clinical presentation and mortality for COVID-19, our model explicitly accounts for such age differences informed by age-specific data. Consequently, our model enables a more granular investigation of disease dynamics and the impact of intervention measures targeting specific age groups. Model predictions of, for example, the time-dependent prevalence in the population can be made for different age groups, which is especially relevant to assess whether herd immunity levels are reached in age groups at the highest risk for severe disease.

Using this model, we evaluated the expected impact of the lockdown and exit strategies for the control of COVID-19 transmission in the population. The basic reproduction number prior to lockdown was estimated to be 2.900 (2.885, 2.918) which is in line with estimates for the epidemic growth in Europe prior to the implementation of nationwide intervention measures and epidemiological modeling in different countries [22, 23, 24, 25, 26, 27], based on recent meta-analytic results [28, 29] and on other modeling exercises specifically tailored to the Belgian setting [30, 31]. Moreover, the intervention measures taken clearly flattened the epidemic curve followed by a progressive reduction of the number of (confirmed) cases over time and the number of new hospitalizations. The decrease in average number of contacts implied a substantial reduction in reproduction number *R*_*t*_ = 0.738 (95% CI: 0.732, 0.744) on May 4, 2020.

The proposed mathematical model is a ‘living’ model used for real-time modeling of the Belgian epidemic and for long-term predictions focusing on, among other things, determining a purchase strategy for medical supplies. Needless to say, the model is updated progressively as new data becomes available and extensions towards improving the model and incorporating up-to-date information are considered in future research. Although several scenarios have been displayed, the single scenario which will unfold in reality in the next weeks and months is driven by unpredictable human behaviour and governmental decisions in case of a resurgence of the disease. Nevertheless, displaying and investigating a range of potential scenarios is crucial in quantifying the impact of certain imposed changes, and of key importance to guide policy makers to shape exit strategies.

Based on the various scenarios presented here, one can conclude that a small to moderate level of transmission in the upcoming months leads to an increased risk of having a large-scale resurgence of the disease later on. In such a situation, a high number of new hospitalizations will be reported with a peak size which is inversely related to the level of sustained transmission in the period preceding the wave of new COVID-19 infections. Lifting the stringent lockdown measures without adequate exit strategies put in place would inevitably have led to a large increase in the number of new infections as the population immunity is still too low to rely on herd immunity (see, e.g., estimated seroprevalence in Figure 5). This signals an insufficient depletion of susceptibles in order to prevent subsequent COVID-19 outbreaks in the future. Our scenario analyses present both short and long-term predictions of new waves based on the current levels of population immunity. However, to date, both the level of protection against infection in the presence of IgG antibodies against SARS-CoV-2 as well as the extent of the (humoral and cellular) immune response in relation to the symptoms of the infected person are still very unclear [5]. In the model, we assume that acquired immunity after recovery lasts for the entire time period under study.

Our work suffers from several limitations. The uncertainty regarding the estimate of the reproduction number on May 4 arises solely from the uncertainty regarding the pre-lockdown reproduction number, given the fact that the uncertainty with regard to the impact of the lockdown (in terms of social contact behavior and non-pharmaceutical interventions) depends on the contact matrix used, hence, this source of variability is absent after selecting the “intervention” contact matrix that provides the best fit to the data. Needless to say, quantification of the effect of the lockdown on the reproduction number compared to the pre-lockdown reproduction number is only possible by assuming the contact behavior prior to the lockdown to be fixed and by having the proportionality factor *q* in the social contact hypothesis to be time-invariant [12]. In our model, the reduction in transmission of COVID-19 is completely attributed to a reduction in social contacts rather than changes in transmissibility due to e.g., use of masks, keeping distance when contacting persons, etc. However, since social contact data collected during the pandemic was unavailable at the start of this project, we were unable to disentangle these effects. A social contact survey (CoMix) done during the lockdown in Belgium measured a reduction of 80% in the overall number of contacts with respect to the pre-lockdown situation [32]. The contact matrix of our best fit model implies an overall median reduction in number of contacts of approximately 75%, so comparable in magnitude. In future work, we will use these social contact data collected in Belgium within the EpiPose project [32] to quantify the impact of the lockdown and its relief on the number of contacts made.

Second, in our model, pre- and asymptomatic individuals on the one hand and persons with mild and severe symptoms (before hospitalization) on the other hand are presumed to have a similar level of infectiousness thereby contributing in the same way to the transmission process. Patients suffering from severe disease probably reduce their contacts more than those with mild disease [33], but the associated reduction in contacts may be compensated by greater infectivity per contact, as more severely affected patients are likely more infectious (i.e., due to a higher viral load) [34]. Moreover the clinical presentation of the disease and disease progression is not uniform with highly variable delay distributions between infection, symptoms, hospitalisation and death. For instance, some individuals with mild symptoms may enjoy a symptom-free intermediate period after which (more severe) symptoms reappear, and immediate hospitalization may be required [35]. In our model, isolation (and treatment) of hospitalized individuals is assumed to lead to a complete reduction in ability to spread the infection. Nevertheless, the contribution of these nosocomial infections is believed to be very limited.

Our model assumes a (potential) differential length of infectiousness between individuals with no symptoms, mild symptoms and those with severe symptoms. This assumption is supported by the faster viral clearance of asymptomatic individuals and individuals with mild symptoms and those individuals with a larger viral load thereby experiencing more severe symptoms [34, 36]. For symptomatic individuals, however, the average duration in the respective I-compartments (*I*_*mild*_ and *I*_*sev*_) before moving to compartment *R* or before being isolated in the hospital (for the severe cases) is a proxy for the (average) duration until individuals completely isolate themselves to prevent subsequent transmission (although they could still be infectious when doing so), rather than being equal to the average infectious period.The correspondence between differential infectious periods depending on symptom severity on the one hand and the serial and generation interval on the other hand is complicated by the fact that the latter quantities depend on both a viral shedding component (linked to infectiousness) and a contact component (which is subject to behavioral change when displaying symptoms) [37]. A theoretical assessment of the link between the serial and generation interval on the one hand and the duration of infectiousness for asymptomatic, mildly infected and severely infected individuals is considered beyond the scope of this manuscript.

The severe compartment in the stochastic model is used as a way to induce a non-exponential delay (generalized Erlang delay distribution) between time of first symptom onset and hospitalization. Faes et al. (2020) [20] showed that the time between symptom onset and hospitalization is indeed non-exponentially distributed, albeit that the best fitting distribution (i.e., a Weibull one) is difficult to incorporate in this modeling framework. As important aspects of the transmission dynamics and the disease are still uncertain, some of the simplifications made in the model will be revisited and updated as biomedical insights improve (e.g., regarding potential seasonality in COVID-19 transmission). As a result of limited information with regard to hospital discharge, the model is currently not able to directly predict the burden on hospital capacity. This will be particularly relevant for surveillance of pressure on the healthcare system in future COVID-19 waves. However, based on the model output in terms of new hospitalizations and information with respect to length of hospital stay, an indirect calculation thereof is straightforward. In the current analyses, we did not distinguish between hospitalization of individuals in elderly homes and individuals from the general population, nor between deaths in hospitals and nursing homes, mainly due to the lack of detailed information to do so. Our model therefore focuses on the general population. Next to that, we disregard potentially important factors such as seasonality (i.e., induced by changes in temperature, humidity, exposure of the virus to ultraviolet light, etc.) entailing an impact on social behaviour and transmission potential of the virus to an extent that is largely unknown to date [38, 39]. Finally, in the scenario analyses presented in this paper, we assume that no external re-importation of the disease in the population occurs, albeit that the stochasticity of the model is able to account for this, at least to a limited extent.

Several mathematical approaches have been considered in the context of the SARS-CoV-2/COVID-19 epidemic in Belgium, all having different merits and limitations [31, 30]. For example, the individual-based model by Willem et al. [31] enabled the direct study of contact tracing and case isolation as a control measure. The meta-population model by Coletti et al. [30] studied the impact of mobility on disease transmission. This stochastic model enabled the detailed fitting to age-specific serology and incidence data using MCMC. As there is no single best model to study all possible research questions related to the spread and control of the disease, we compared model outputs and conclusions, as their predictions need continuous finetuning and validation [40, 41]. In conclusion, predictions from our model are useful to inform subsequent serological sample studies and to explore various exit scenarios with respect to disease transmission as an input for the investigation of the economic impact of COVID-19 epidemics on society.

## Data Availability

Data on hospitalizations and deaths are available on the website of the Belgian Scientific Institute for Public Health, Sciensano.

https://epistat.wiv-isp.be/Covid/

## 5 Acknowledgements

We thank several researchers from the SIMID COVID-19 consortium from the University of Antwerp and Hasselt University for numerous constructive discussions and meetings. SA and NH gratefully acknowledge support from the Research Foundation Flanders (FWO) (RESTORE project – G0G2920N). This work also received funding from the European Research Council (ERC) under the European Union’s Horizon 2020 research and innovation program (PC and NH, grant number 682540 – TransMID project, NH, PB grant number 101003688 – EpiPose project). The authors are also very grateful for access to the data from the Belgian Scientific Institute for Public Health, Sciensano, and from the Vaccine & Infectious Disease Institute (VaxInfectio), University of Antwerp. LW and PL gratefully acknowledge funding from the Research Foundation Flanders (postdoctoral fellowships 1234620N and 1242021N). We acknowledge support from the Antwerp Study Centre for Infectious Diseases (ASCID). The resources and services used in this work were provided by the VSC (Flemish Supercomputer Center), funded by the FWO and the Flemish Government. The funders had no role in study design, data collection and analysis, decision to publish, or preparation of the manuscript.

## 6 Author contributions

SA, JW and NH conceived the study. SA and JW contributed to the software development. SA, PC, LW, PB and NH prepared the first draft of the manuscript. SA, PC, CF, SH, SM, OP and JW contributed to the data preparation and/or collection. All authors contributed to the final version of the paper and approved the final version of the manuscript.

## A Terminology

In Table A1, we present an overview of the different states and the terminology used throughout the manuscript.

**Table A1:** Notations related to the compartments in the model

| State | Description |
| --- | --- |
| $S(t)$ | Susceptible individuals |
| $E(t)$ | Infected individuals not being infectious yet |
| $I_{presym}(t)$ | Individuals pre-symptomatically infected |
| $I_{asym}(t)$ | Asymptomatically infected individuals |
| $I_{mild}(t)$ | Infected individuals with mild symptoms |
| $I_{sev}(t)$ | Infected individuals with severe symptoms requiring hospitalization |
| $I_{hosp}(t)$ | Hospitalized individuals |
| $I_{icu}(t)$ | Individuals admitted to intensive care unit |
| $D(t)$ | Number of deaths |
| $R(t)$ | Recovered individuals |

## B Model parameters

In this appendix, we provide a detailed overview of the different assumptions related to the transmission process and the change in behaviour upon (severe) symptomatic infection.

### B.1 Susceptibility and infectiousness in children

Until recently, there was no (conclusive) evidence of differential infectiousness and/or susceptibility to COVID-19 infection in children. Some authors claimed that no significant differences in viral load were present between (symptomatic) children and adults, even after revising their work [1]. A re-analysis of these (initial) findings by Held and McConway and Spiegelhalter [2, 3], however, clearly shows that children between 1 and 10 years old have on average only 27% (95% CI: 8% - 91%) of the viral load of adults aged 20 years or more. For the mathematical model proposed here, we do not differentiate between infectiousness and susceptibility in children as compared to adults directly [4]. However, as children are presumed to be more likely to have a higher probability of being asymptomatically infected (see Section B.2), and the relative infectiousness of asymptomatic versus symptomatic individuals *r*_*β*_ is (assumed) equal to 0.51 [5], children are implicitly less infectious, hence, contribute less to the transmission process relative to adults.

### B.2 Age-dependent proportions of asymptomatic cases

The age-dependent proportions of asymptomatic cases, represented by the vector ***p***, are based on a study by Wu et al. (2020) [6]. More specifically, we use the age-specific relative susceptibility to symptomatic infection reported by Wu et al. (2020) [6] to inform the proportion of asymptomatic cases. In order to do so, we start from an overall age-weighted proportion of asymptomatic cases in the Belgian population equal to 50% [6] implying that

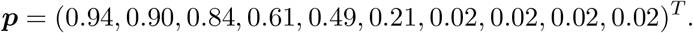

Although we are fixing the age-dependent proportions of asymptomatic cases in the model, we do allow for differential probabilities of hospitalization through the specification of an age-dependent probability of only having mild symptoms upon being symptomatic. The reason for this constraint is the fact that based on the available data, we cannot disentangle the age-specific probability of being asymptomatic from the probability of having mild symptoms.

**Figure B12:**
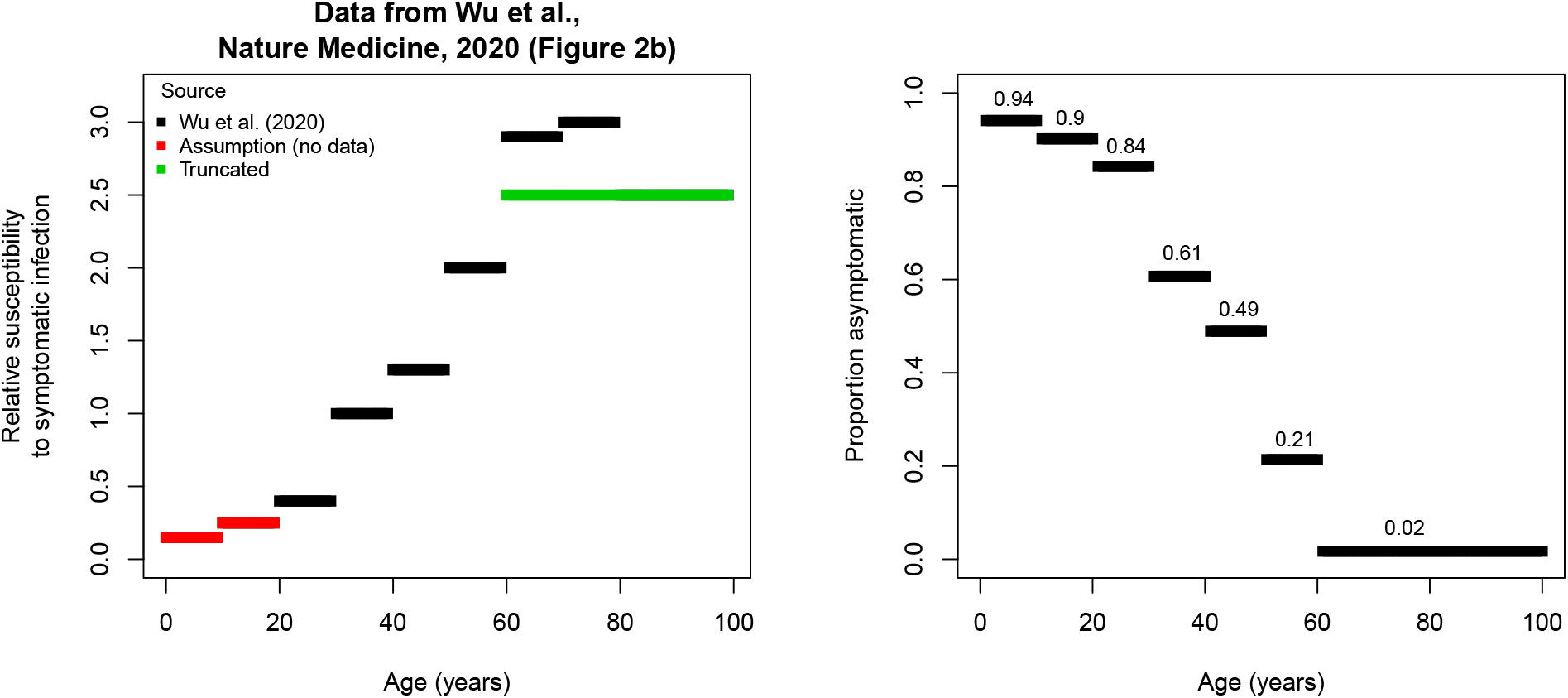
Relative susceptibility to symptomatic infection as a function of age (in years) reported by Wu et al. (2020) [6] (left panel) and the proportion of asymptomatic cases by age relying on the assumption of 50% of asymptomatic infections.

### Symptom severity upon symptomatic infection

In Table B1, the proportion of hospitalizations and ICU admissions are reported for COVID-19 cases by age group in the United States (between February 12 - March 16, 2020) [7].

**Table B1:** Hospitalization and Intensive Care Unit (ICU) admission percentages (%) for reported COVID-19 cases by age group based on data from the USA, February 12 - March 16, 2020 [7].

| Age group | Hospitalization | Average | ICU | Average | Age group | $\phi_0$ | $\phi_1$ |
| --- | --- | --- | --- | --- | --- | --- | --- |
| 0-19 | 1.6-2.5 | 2.05 | 0.0 | 0.00 | [0,10) | 98.0 | 100.0 |
| 20-44 | 14.3-20.8 | 17.55 | 2.0-4.2 | 3.10 | [10,20) | 98.0 | 100.0 |
| 45-54 | 21.2-28.3 | 24.75 | 5.4-10.4 | 7.90 | [20,30) | 79.0 | 85.0 |
| 55-64 | 20.5-30.1 | 25.30 | 4.7-11.2 | 7.95 | [30,40) | 79.0 | 85.0 |
| 65-74 | 28.6-43.5 | 36.05 | 8.1-18.8 | 13.45 | [40,50) | 67.0 | 76.0 |
| 75-84 | 30.5-58.7 | 44.60 | 10.5-30.0 | 20.25 | [50,60) | 67.0 | 76.0 |
| $\geq 85$ | 31.3-70.3 | 50.80 | 6.3-29.0 | 17.65 | [60,70) | 50.0 | 73.0 |
|  |  |  |  |  | [70,80) | 35.0 | 69.0 |
| Total | 20.7-31.4 | 26.05 | 4.9-11.5 | 8.20 | [80,100) | 32.0 | 74.0 |

In our model, we estimate the age-specific probability of developing only mild symptoms ***ϕ***_0_. Furthermore, the probabilities ***ϕ***_1_ are fixed to the values reported in Table B1. Note that if, for example, *ϕ*_1_(*k*) = 0.75 and *ϕ*_0_(*k*) = 0.8, representing the probability of hospitalization conditional on having severe symptoms and the probability of having mild symptoms in age group *k*, then the probability of hospitalization in symptomatic individuals is equal to 0.2 × 0.75 = 0.15. Since we lack detailed age-specific information about the relative proportions of ICU admissions as compared to the total number of hospitalizations, and data on referral between ICU and hospital wards throughout hospital stay, we are not able to inform ***ϕ***_1_.

### B.4 Case fatality rates - Probability of dying upon hospitalization

Age-dependent case fatality rates have been adopted from Riou et al. (2020) [8] which were estimated from outbreak data obtained in Hubei, China in the period January to February. In Table B2, we present these age-specific case-fatality rates 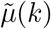 (number of deaths relative to confirmed cases). In our analysis, the rates *µ*(*k*) are rather representing the probability of dying upon hospitalization (deaths relative to number of hospitalized individuals), taken to be equal for hospitalized individuals and critically ill individuals (i.e., *µ*(*k*) *≡ µ*_*hosp*_(*k*) = *µ*_*icu*_(*k*)), which also relates to the inability of disentangling hospitalization from ICU admission based on the available data. In the stochastic model, however, we estimate these rates *µ*(*k*) from the available mortality data. Due to the low number of deaths in young age categories, we assume that *µ*(1) = 0 and *µ*(2) = *µ*(3), comprising parameters to be estimated from the data. A complete list of model parameters with reference values is presented in Table B3.

**Table B2:** Age-dependent case-fatality rates.

| [0-10) | [10-20) | [20-30) | [30-40) | [40-50) | [50-60) | [60-70) | [70-80) | [80-90) | [90, ∞) |
| --- | --- | --- | --- | --- | --- | --- | --- | --- | --- |
| 0.000094 | 0.00022 | 0.00091 | 0.0018 | 0.004 | 0.013 | 0.046 | 0.098 | 0.18 | 0.18 |

### B.5 Overview of epidemiological and model parameters

An overview of important epidemiological parameters related to SARS-CoV-2/COVID-19 transmission dynamics is provided in Table B3 together with relevant sources. Some of these parameters are directly or indirectly included in the modeling approach. In the last column we indicate whether these parameters are estimated or fixed in the estimation procedure. A detailed overview of all model parameters can be found in the next subsection.

### B.6 Prior distributions

In Table B4, we present an overview of the prior distributions considered for the various model parameters.

**Table B3:** List of epidemiological parameters; ^*^: for asymptomatic individuals only.

| Notation | Description | Value | Source | Model |
| --- | --- | --- | --- | --- |
| $\rho^{-1}$ | Average length of incubation period | 5.2 (4.1, 7.0)<br>5.2 days<br>5 days<br>4 days | [5]<br>[9]<br>[10]<br>[11] | Estimated |
| $\gamma^{-1}$ | Average length of latency period | 2 days | – | Estimated |
| $\theta^{-1}$ | Average length of pre-symptomatic infectious period | 3.2 days | $\rho^{-1} - \gamma^{-1}$ | Estimated |
| $\sigma^{-1}$ | Average length of infectious period | 4.5 days<br>5 days*<br>6 days*<br>5–7 days<br>9.3 (7.8, 10.0) | [12]<br>[13]<br>[14]<br>[4, 15]<br>[16] | – |
| $\omega^{-1} + \delta_2^{*-1}$ | Average time between (severe) symptom onset and hospitalization | 5.9 (0.8, 11.0)<br>2.7 (1.6, 4.1)<br>3 – 10.4 | [17]<br>[10]<br>[18] | Estimated |
| $\delta_1^{-1}$ | Average length of infectious period when asymptotically infected (after pre-symptomatic phase) | 3.5 days | – | Estimated |
| $\delta_2^{*-1}$ | Average length of infectious period when mildly infected (after pre-symptomatic phase) | 3.5 days | – | Estimated |
| $\delta_3^{*-1}$ | Recovery period of hospitalized individuals – length of stay in the hospital | 11.5 (8.0, 17.3)<br>13.3 (7.3, 19.3) | [19]<br>[17] | Estimated |
| $\delta_4^{*-1}$ | Recovery period of critically infected in ICU – length of stay in ICU | 14 days | – | Estimated |
| $p$ | Proportion of asymptomatic cases | 0.42 (0.17, 0.67)<br>0.35 (0.30, 0.39) | [20, 21]<br>[22] | Fixed |
| $1 - \phi_0$ | Proportion of symptomatic cases developing severe to very severe symptoms | 0 (children)<br>0.1 (adults)<br>0.2 (seniors)<br>0.2 (constant) | [23]<br>[24]<br>[25]<br>– | Estimated |
| $\phi_0$ | Proportion of symptomatic cases developing only mild symptoms | $1 - (1 - \phi_0)$ | – | Estimated |
| $1 - \phi_1$ | Proportion of hospitalized persons requiring ICU admission | 0.25 (constant) | – | Fixed |
| $r_\beta$ | Relative infectiousness of pre-symptomatic and asymptomatic individuals | 0.51 | [5] | Fixed |
| $R_0$ | Basic reproduction number | 2.24 (1.96, 2.25)<br>2.35 (1.15, 4.77)<br>2.36, 2.5<br>3.00 (2.80, 3.20)<br>3.00 (2.60, 3.30)<br>3.10 (2.90, 3.20)<br>3.60 (3.49, 3.84)<br>3.60 (3.20, 4.20) | [26]<br>[27]<br>[28, 29]<br>[30]<br>[31]<br>[32]<br>[33]<br>[34] | Estimated |

**Table B4:** Prior distributions for the model parameters.

| Parameter | Baseline scenario |  |
| --- | --- | --- |
|  | Prior distribution | # parameters |
| $\gamma$ | $N(0.500, 0.050^2)$ | 1 |
| $\theta$ | $N(0.286, 0.050^2)$ | 1 |
| $\delta_1$ | $N(0.286, 0.050^2)$ | 1 |
| $\delta_2^*$ | $\text{Un}(0, 1)$ | 1 |
| $\delta_3^*$ | $\text{Un}(0, 1)$ | 1 |
| $\omega(k)$ | $\text{Un}(0, 1)$ | 10 |
| $\beta_1^*$ | $\text{Un}(0, 5)$ | 1 |
| $\phi_0(k)$ | $\text{Un}(0, 1)$ | 10 |
| $\mu(k)$ | $\text{Un}(0, 1)$ | 8 |
| $R_0$ | $N(2.500, 0.100^2)$ | 1 |

## C Social contact data

### C.1 Baseline contact matrices

The contact matrices for the asymptomatic individuals (***C***_*asym*_) is taken to be equal to the general contact matrices collected in the Belgian survey in 2010–2011. Thus we assume that individuals do not change their contact behaviour when being asymptomatically infected with COVID-19. The overall contact matrix is the sum of the contact matrices encompassing contacts made at the following locations: home, work, school, transportation, leisure and other places.

Thus ***C***_*asym*_ is obtained as follows:

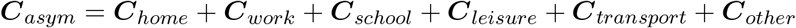

In Figure C1, we graphically depict the social contact matrix ***C***_*asym*_ in terms of the average number of daily contacts for individuals of different age groups contacting each other. The contact matrices for symptomatic individuals are obtained by re-scaling the matrix ***C***_*asym*_ in the respective locations by the relative change in the number of contacts observed by Van Kerckhove et al. [35] during the 2009 A/H1N1 pandemic Influenza in England. Hence, we presume that social contacts are adapted in a similar way in the Belgian population upon contracting the disease and experiencing symptoms. Thus, ***C***_*sym*_ is defined as a weighted sum of the aforementioned contact matrices at specific locations, i.e.,

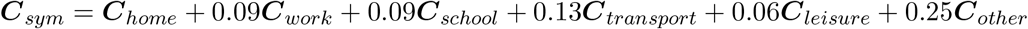

### C.2 Intervention contact matrices

The contact matrices for the asymptomatic individuals during the lockdown depend on the intervention considered (see Table 1). The contact matrices made in all locations are changed except for the one accommodating contacts made at home. The framework of assigning relative reductions to the social contact matrices obtained at the different locations can be illustrated as follows:

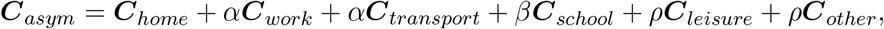

where 1 − *α* represents the percentage of telework considered (people working from home and/or who have stopped working), *β* represents the percentage of school contacts retained (hence, 0 in case of school closure), *ρ* represents the fraction of contacts during leisure and other activities that are still made given the imposed measures targeting physical distancing. The contact matrices of the symptomatic ***C***_*sym*_ are obtained in a similar manner:

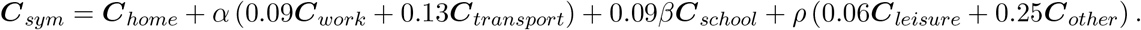

**Figure C1:**
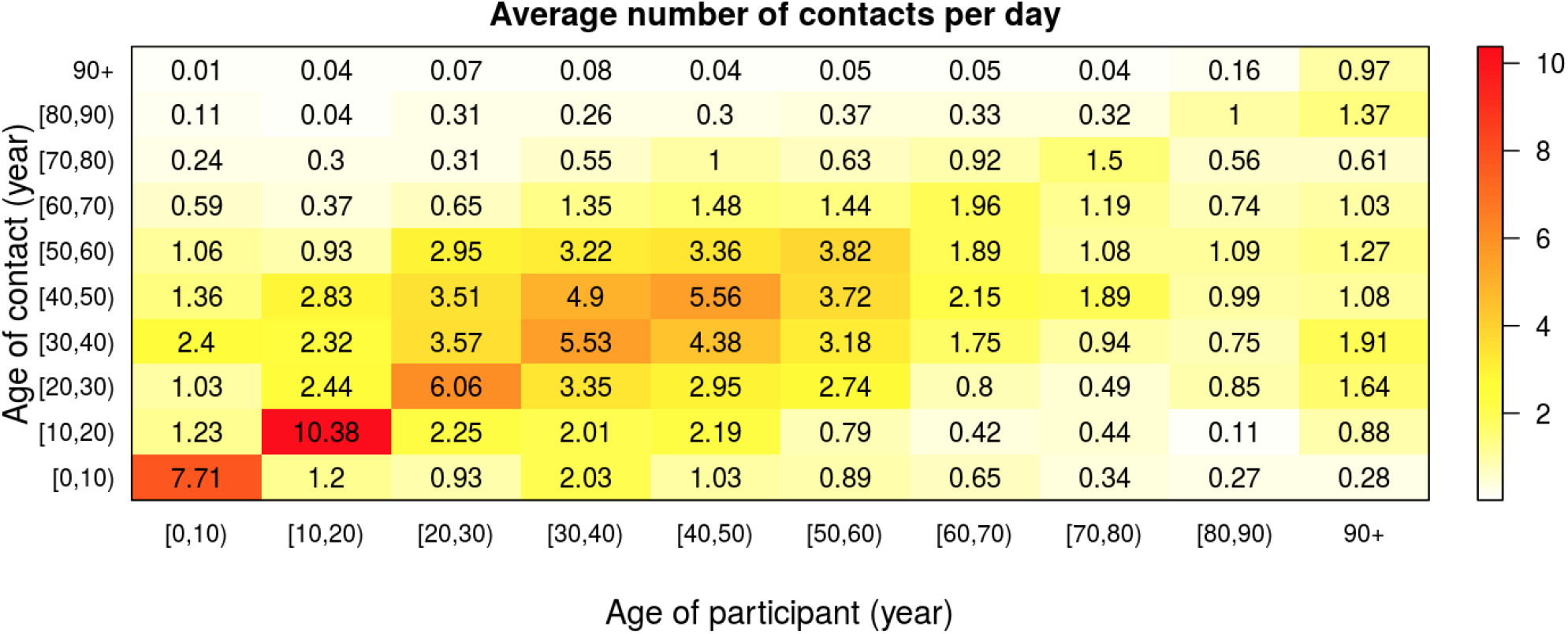
Average number of contacts per day between individuals of different age classes - social contact matrix ***C***_***asym***_ based on the social contact data from Belgium anno 2010–2011.

All location-specific contact matrices from the Belgian social contact survey in 2010 are directly available at http://www.socialcontactdata.org/socrates/.

**Figure C2:**
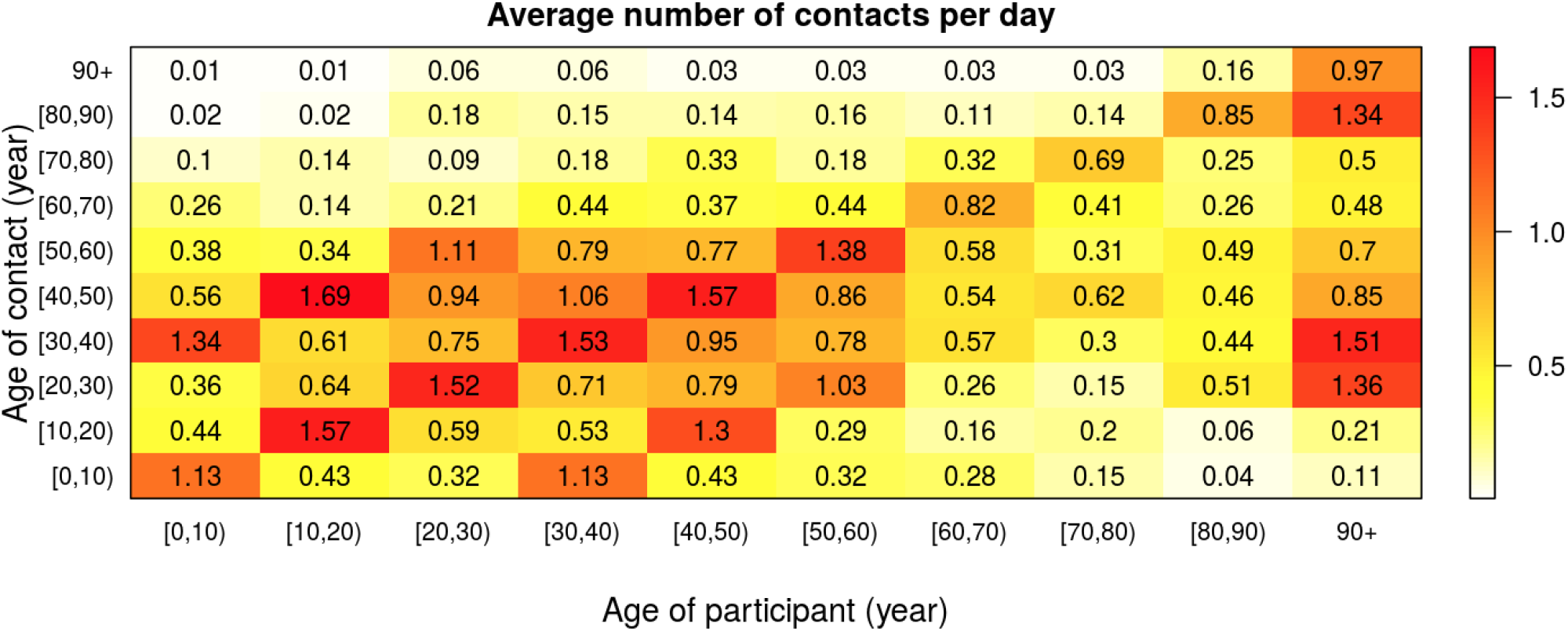
Average number of contacts per day between individuals of different age classes - social contact matrix ***C***_***asym***_ based on the social contact data from Belgium anno 2010–2011 after intervention measures are imposed according to the 80% TW & SC scenario outlined in Table 1.

The performance of the matrices presented in Table 1 are compared based on the Deviance Information Criterion (DIC), introduced by Spiegelhalter et al. [36] to compare the relative fit of a set of Bayesian hierarchical models. DIC is a relative measure balancing goodness-of-fit and complexity of a model and is based on the deviance. An overview of the respective DIC-values related to the different choices of the intervention contact matrices is presented in Table C5.

**Table C5:** Deviance Information Criterion (DIC) values for the different social contact matrices considered to quantify the impact of the intervention measures on social contact patterns. Percentage of average number of pre-pandemic contacts at different locations. WT: Work and transport reductions, SC: School closure.

| Social contact matrix | Work & Transport | School closure | Leisure & other activities | DIC |
| --- | --- | --- | --- | --- |
| 50% WT & SC | 50% | Yes | 10% | 12930.98 |
| 60% WT & SC | 40% | Yes | 10% | 12896.75 |
| 70% WT & SC | 30% | Yes | 10% | 12459.82 |
| 80% WT & SC | 20% | Yes | 10% | 11577.97 |
| 90% WT & SC | 10% | Yes | 10% | 12477.25 |

## D Hospitalization incidence data

In Figure D1, we depict the weekly age distribution of hospitalized cases derived from the clinical surveillance database of COVID-19 hospitalized patients during the first wave of COVID-19 in Belgium. As mentioned in the main text, the day of introduction is considered to be March 1, 2020.

**Figure D1:**
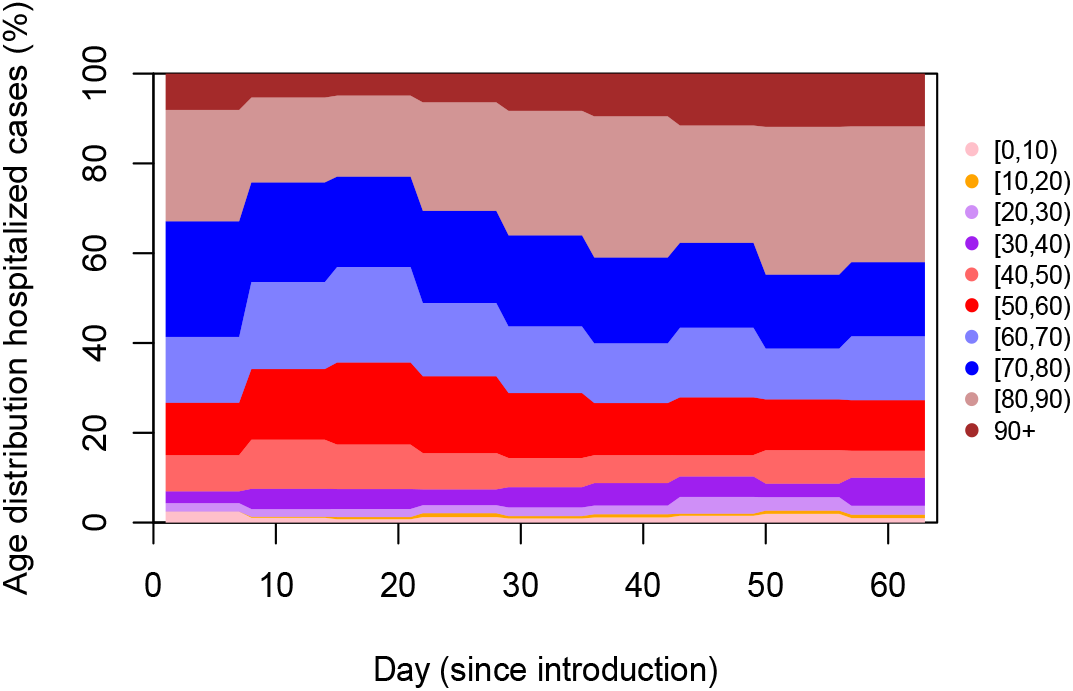
Weekly age distribution of hospitalized patients in Belgian hospitals during the first COVID-19 wave in Belgium.

## E Serological survey data

We use serial serological survey data collected during two cross-sectional periods and based on residual samples coming from 10 private laboratories [37]. Serological survey data is collected at different cross-sectional sampling times and blood samples are tested for the presence of IgG antibodies against the SARS-CoV-2 virus. Consequently, individuals are classified as seronegative, equivocal or seropositive based on their measured IgG antibody concentration against S1 proteins of SARS-CoV-2 obtained from the EuroImmun semi-quantitative ELISA test kit (EuroImmun, Luebeck, Germany). The age-specific seroprevalence is derived as the proportion of seropositive individuals in each age class.

In order to relate the model-based prevalence of COVID-19 in the population to the observed seroprevalence, we assume that the seroprevalence at calendar time *t* and age *a* is denoted by *π*(*a, t*) and that IgG antibodies against SARS-CoV-2 are detectable upon infection according to the following logistic function:

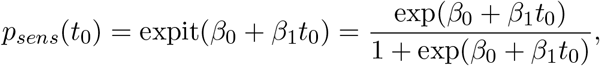

where *p*_*sens*_(*t*_0_) represents the probability of having a sufficiently high IgG antibody concentration to indicate past SARS-CoV-2 infection. The sensitivity of the diagnostic tests is considered to be a function of the time since symptom onset *t*_0_ (at least in the presence of symptoms), i.e. sensitivity of the diagnostic testing procedure as a function of time since onset of symptoms presuming a sensitivity of zero prior to symptom onset. In this modeling approach, we rely on estimates of the sensitivity curve obtained from the literature [38], reaching a sensitivity of 99% 14 days after symptom onset, albeit that sufficient information regarding the sensitivity of the specific diagnostic test in use is currently lacking. In this exercise, specificity of the test is presumed to be very high (100%), implying no false positive test results. A lower specificity would lead to more false positive cases, thereby overestimating the seroprevalence as compared to the true underlying prevalence in the population. Since the model is calibrated on hospitalization data, the increase in false positives would imply an underestimation of the probability of hospitalization and an overestimation of the total number of infected cases in the population (the so-called dark number). However, we do believe that the general conclusions with regard to the impact of exit strategies on the burden of the healthcare system through the number of hospitalizations and deaths are not affected by this lower specificity, especially since there is little acquired immunity.

The total number of individuals of age *a* at the time of data collection *t* (expressed as days since the start of the epidemic) in the population that will test positive can be written as:

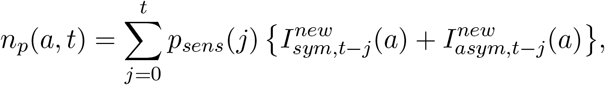

where 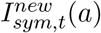 and 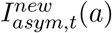 refer to the number of new individuals of age *a* with symptom onset at time *t* and the number of individuals of age *a* entering the asymptomatic state at time *t*, respectively. Although still uncertain to date, asymptomatic individuals are presumed to be similar to symptomatic ones in terms of their humoral immune response following exposure to the SARS-CoV-2 virus.

More recently, Borremans et al. (2020) [39] showed that IgG antibody detection probabilities increase with time since symptom onset, implying that nearly all (98–100% of) individuals had detectable antibodies by day 22–23 after symptom onset. Although detection probabilities are estimated based on different assays, these authors showed that all assays exhibit comparable growth rates except for a slower increase in antibody levels for IgG ELISA-Spike assays. As a sensitivity analysis, we show the impact of altering the logistic sensitivity curve *p*_*sens*_(*t*_0_) (black line) with a delayed 99% detectability of IgG antibodies in line with the aforementioned findings by Borremans et al. (2020) (red line in Figure E2) in Appendix F.

**Figure E2:**
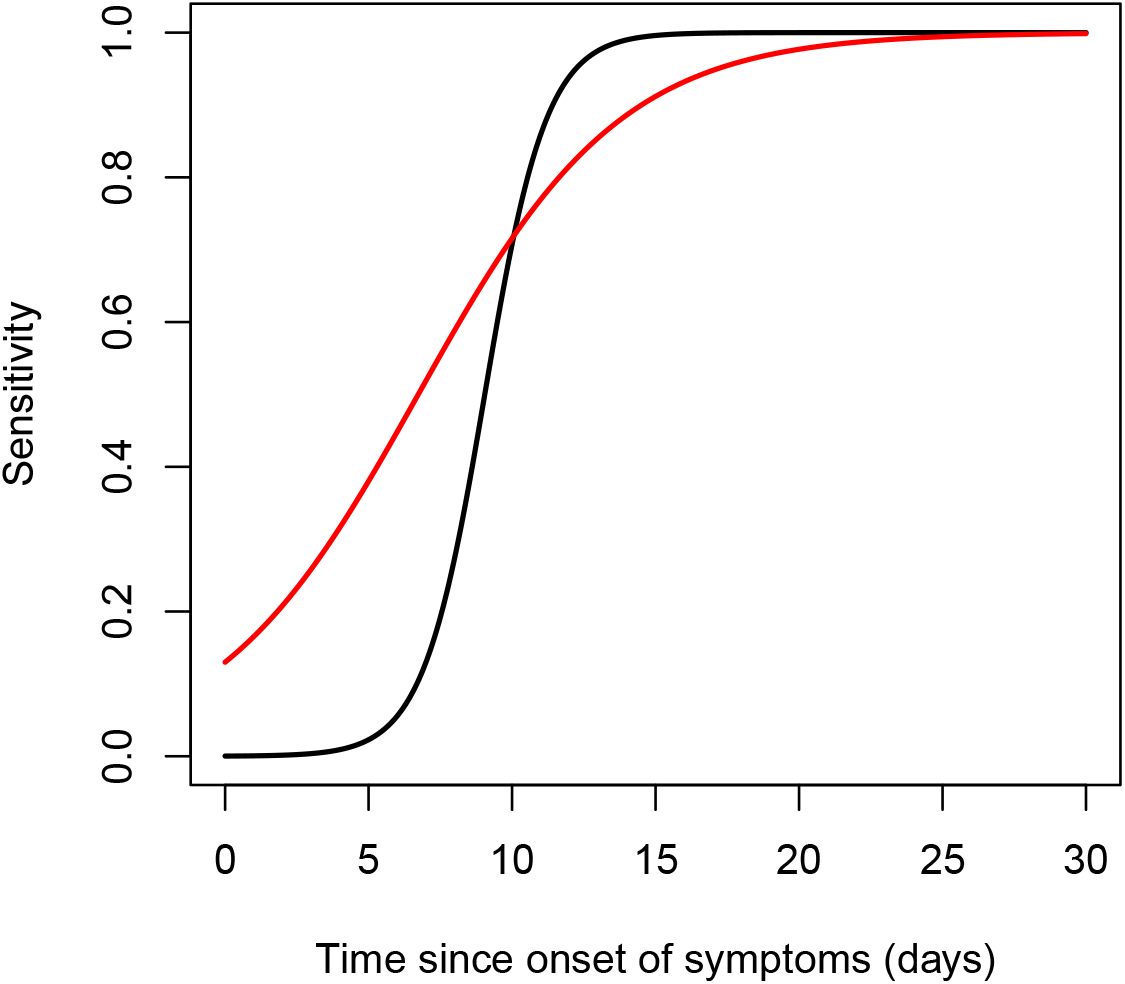
Presumed sensitivity curves with time since onset of symptoms. The black solid line represents the sensitivity curve constructed based on results in Lou et al. (2020) [38] and the red line is based on the findings by Borremans et al. (2020) [39].

## F Additional results

### F.1 Posterior summary measures

Here, we present an overview of the posterior mean, median and 95% credible intervals (CIs) for all model parameters, and parameters derived thereof, in the final model (Table F1). In total, the final model has 35 parameters. As mentioned in Appendix B, the vectors defining the age-specific probability of being asymptomatic ***p*** and the probability of regular hospitalization versus ICU admission ***ϕ***_1_ are fixed, 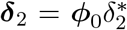 and 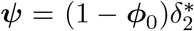. Moreover, 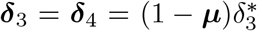 and 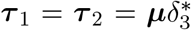. Note that *µ*(1) is fixed to zero as there are no deaths observed in the age group [0, 10) and *µ*(2) = *µ*(3) given the very small number of deaths in age groups [10, 20) and [20, 30). Furthermore, *q* is an implicit model parameter governing the extent of *R*_0_. Prior distributions for all 35 model parameters are listed in Table B4.

### F.2 Number of individuals in different compartments

In Figure F1, the evolution of the proportions of susceptible, exposed, pre-symptomatic, asymptomatic, mildly infected and individuals with severe symptoms (prior to hospitalization) are shown by age group until May 4, 2020.

#### F.3 Compliance to intervention measures

The compliance to the intervention measures is modelled using a logistic curve which is depicted in Figure F2. Full compliance to the measures is reached after approximately 6 days.

**Table F1:** Posterior mean, median and 95% credible interval for the model parameters; CI: credible interval.

| Parameter | Mean | Median | 95% CI | Parameter | Mean | Median | 95% CI |
| --- | --- | --- | --- | --- | --- | --- | --- |
| $q$ | 0.051 | 0.051 | (0.049, 0.54) | $\delta_1$ | 0.240 | 0.240 | (0.224, 0.262) |
| $\gamma$ | 0.729 | 0.730 | (0.672, 0.791) | $\delta_2^*$ | 0.756 | 0.756 | (0.713, 0.806) |
| $\theta$ | 0.475 | 0.476 | (0.434, 0.523) | $\delta_3^*$ | 0.185 | 0.185 | (0.171, 0.200) |
| $\beta_1^*$ | 1.404 | 1.401 | (1.283, 1.551) | $R_0$ | 2.900 | 2.899 | (2.885, 2.918) |
| $\phi_0(1)$ | 0.972 | 0.972 | (0.968, 0.975) | $\phi_0(6)$ | 0.971 | 0.971 | (0.969, 0.972) |
| $\phi_0(2)$ | 0.992 | 0.992 | (0.991, 0.993) | $\phi_0(7)$ | 0.958 | 0.958 | (0.956, 0.960) |
| $\phi_0(3)$ | 0.984 | 0.984 | (0.982, 0.985) | $\phi_0(8)$ | 0.926 | 0.926 | (0.922, 0.929) |
| $\phi_0(4)$ | 0.987 | 0.987 | (0.986, 0.988) | $\phi_0(9)$ | 0.956 | 0.956 | (0.954, 0.958) |
| $\phi_0(5)$ | 0.977 | 0.977 | (0.975, 0.978) | $\phi_0(10)$ | 0.926 | 0.926 | (0.921, 0.930) |
| $\omega(1)$ | 0.167 | 0.142 | (0.102, 0.335) | $\omega(6)$ | 0.275 | 0.275 | (0.244, 0.306) |
| $\omega(2)$ | 0.095 | 0.094 | (0.061, 0.139) | $\omega(7)$ | 0.343 | 0.342 | (0.315, 0.371) |
| $\omega(3)$ | 0.099 | 0.099 | (0.080, 0.120) | $\omega(8)$ | 0.378 | 0.374 | (0.342, 0.431) |
| $\omega(4)$ | 0.162 | 0.161 | (0.142, 0.187) | $\omega(9)$ | 0.334 | 0.335 | (0.308, 0.361) |
| $\omega(5)$ | 0.338 | 0.338 | (0.306, 0.370) | $\omega(10)$ | 0.302 | 0.304 | (0.252, 0.336) |
| $\mu(1)$ | — | — | — | $\mu(6)$ | 0.068 | 0.068 | (0.060, 0.076) |
| $\mu(2)$ | 0.005 | 0.005 | (0.001, 0.011) | $\mu(7)$ | 0.183 | 0.184 | (0.170, 0.195) |
| $\mu(3)$ | 0.005 | 0.005 | (0.001, 0.011) | $\mu(8)$ | 0.325 | 0.325 | (0.310, 0.340) |
| $\mu(4)$ | 0.024 | 0.024 | (0.017, 0.033) | $\mu(9)$ | 0.446 | 0.446 | (0.430, 0.464) |
| $\mu(5)$ | 0.037 | 0.037 | (0.030, 0.045) | $\mu(10)$ | 0.611 | 0.611 | (0.568, 0.653) |

### F.4 Time-dependent reproduction number

In Figure F3, we display the effective reproduction number over time with a 95% credible interval as a red shaded area around the posterior mean. The reproduction number decreased from 2.900 prior to the lockdown to a value of 0.738 (95% CI: 0.732, 0.744) on May 4, 2020.

### F.5 Mortality rates

Age-specific mortality rates are presented in Figure F4. Mortality rates are considered to be equal for persons admitted to ICU and to a regular hospital ward as no distinction could be made between referrals within hospitals, nor between deaths in ICU and hospital wards. Needless to say, mortality rates increase by age with the highest mortality rate in the 90+ age category.

### F.6 Infection fatality rates

Infection fatality rates (IFRs) are calculated based on the observed number of deaths by age group and the estimated total number of infections in a specific age group (thereby accounting for asymptomatic infections) by May 4, 2020. The overall IFR is estimated to be equal to 0.507% (95% credible interval: 0.480%, 0.536%), excluding nursing home deaths. Note that the model disregards the number of deaths in elderly homes (as this is considered to be a separate COVID-19 outbreak which is not fully accommodated in the considered approach), thereby potentially underestimating the IFRs in the highest age groups, at least if the increase in deaths exceeds the increase in additional infections in nursing homes. Next to that, individuals infected prior to May 4, 2020 that will pass away after that date are not included in the calculations, thereby underestimating the IFRs. A more thorough estimation of the IFRs is performed by Molenberghs et al. (2020) [40], accommodating delay in mortality following infection. According to Molenberghs et al. (2020), the IFRs in the nursing home population are much higher compared to those in the non-nursing home population and differences in IFRs in older age categories are linked to frailty and underlying prevalence of comorbidities, characteristics which are very much different in nursing and non-nursing home populations.

**Figure F1:**
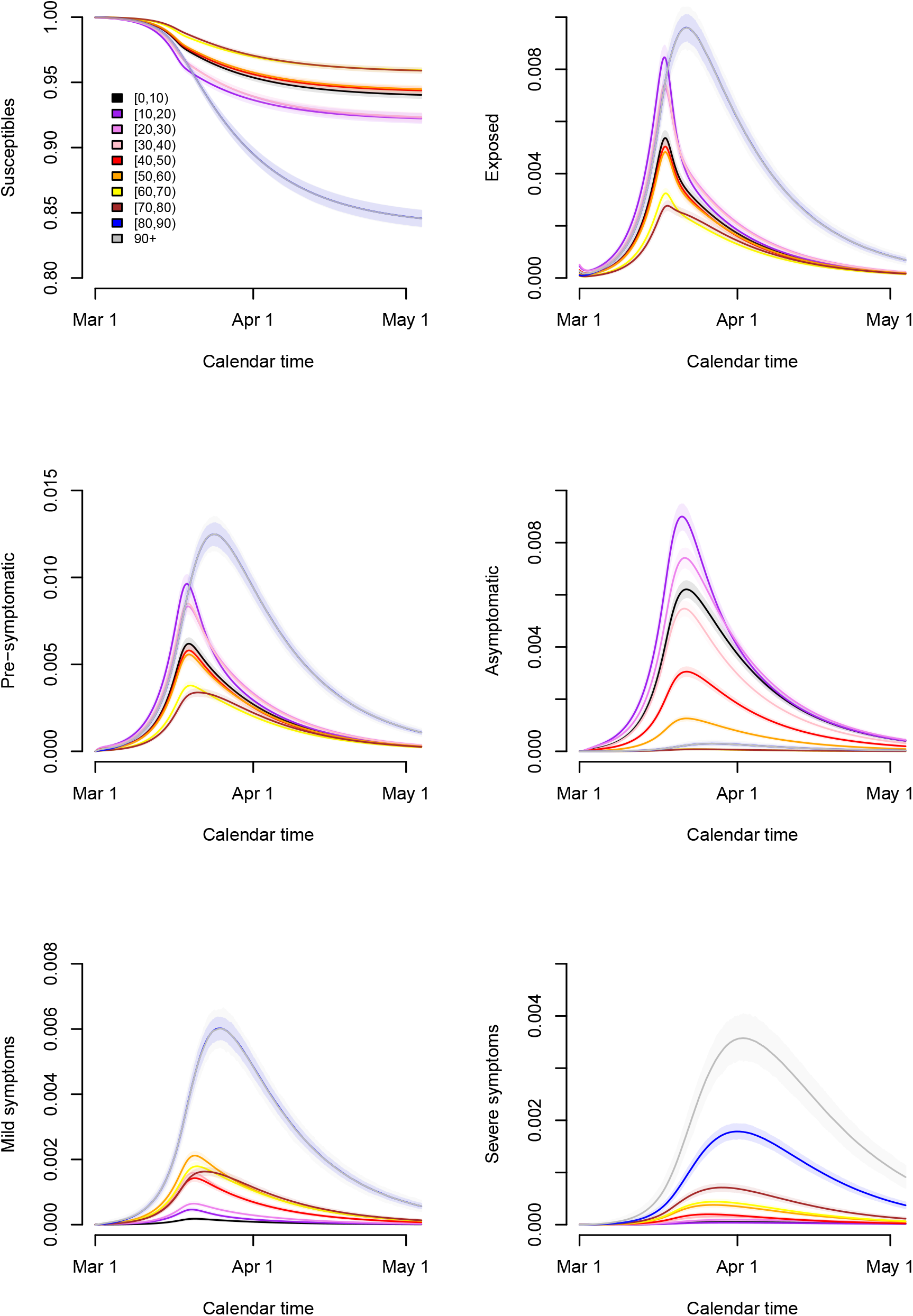
Evolution of the proportion of individuals in the different compartments by age group.

**Figure F2:**
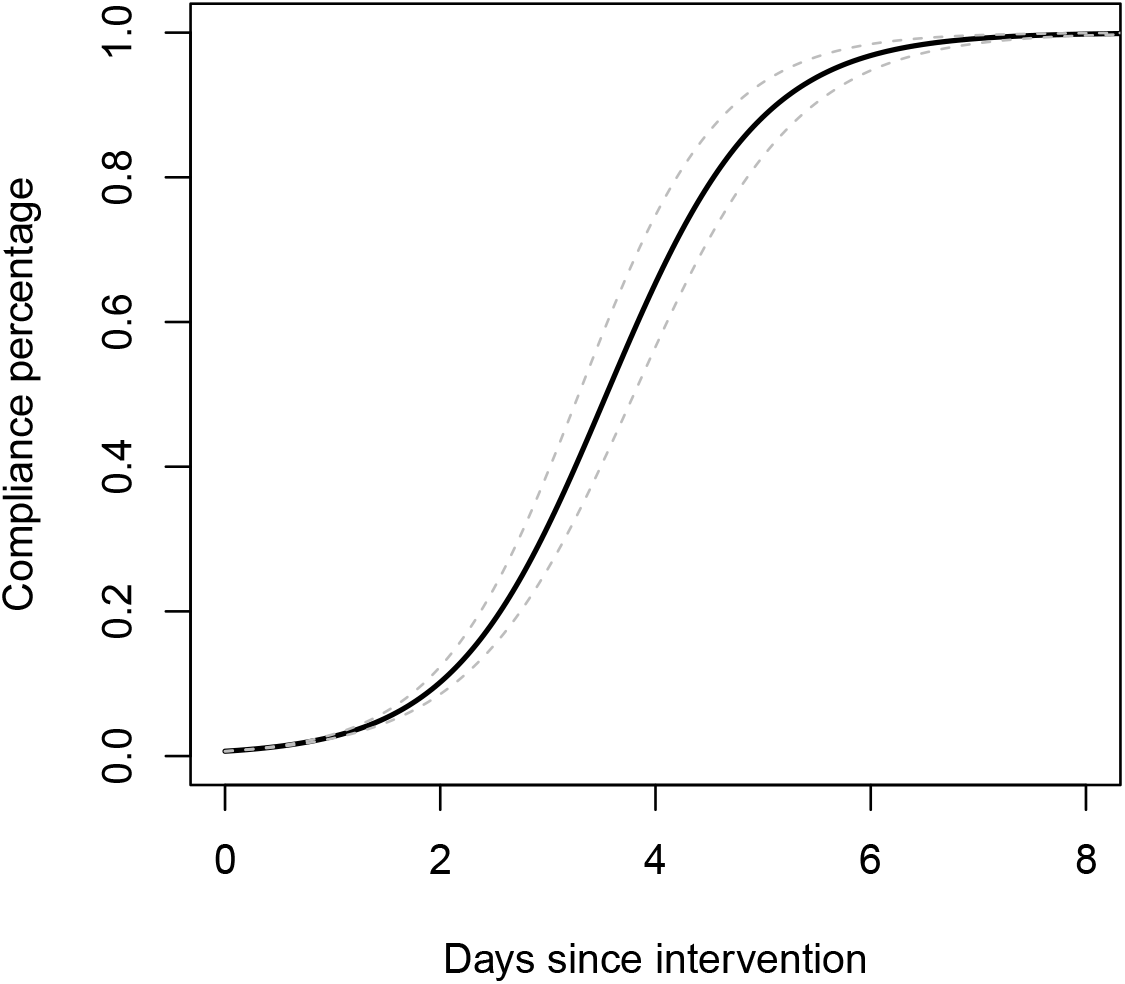
Estimated compliance function to the intervention measures taken by the government.

**Figure F3:**
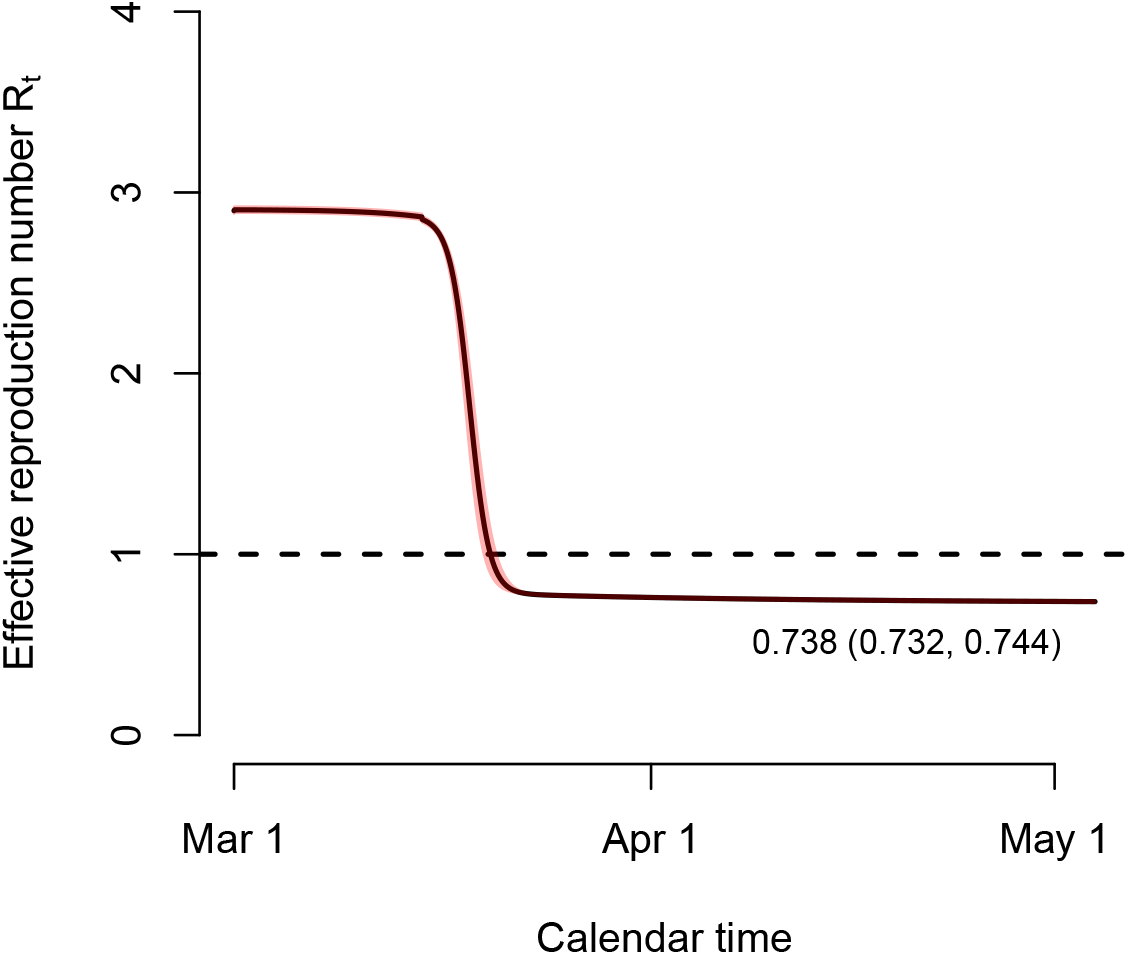
Time-dependent effective reproduction number.

**Figure F4:**
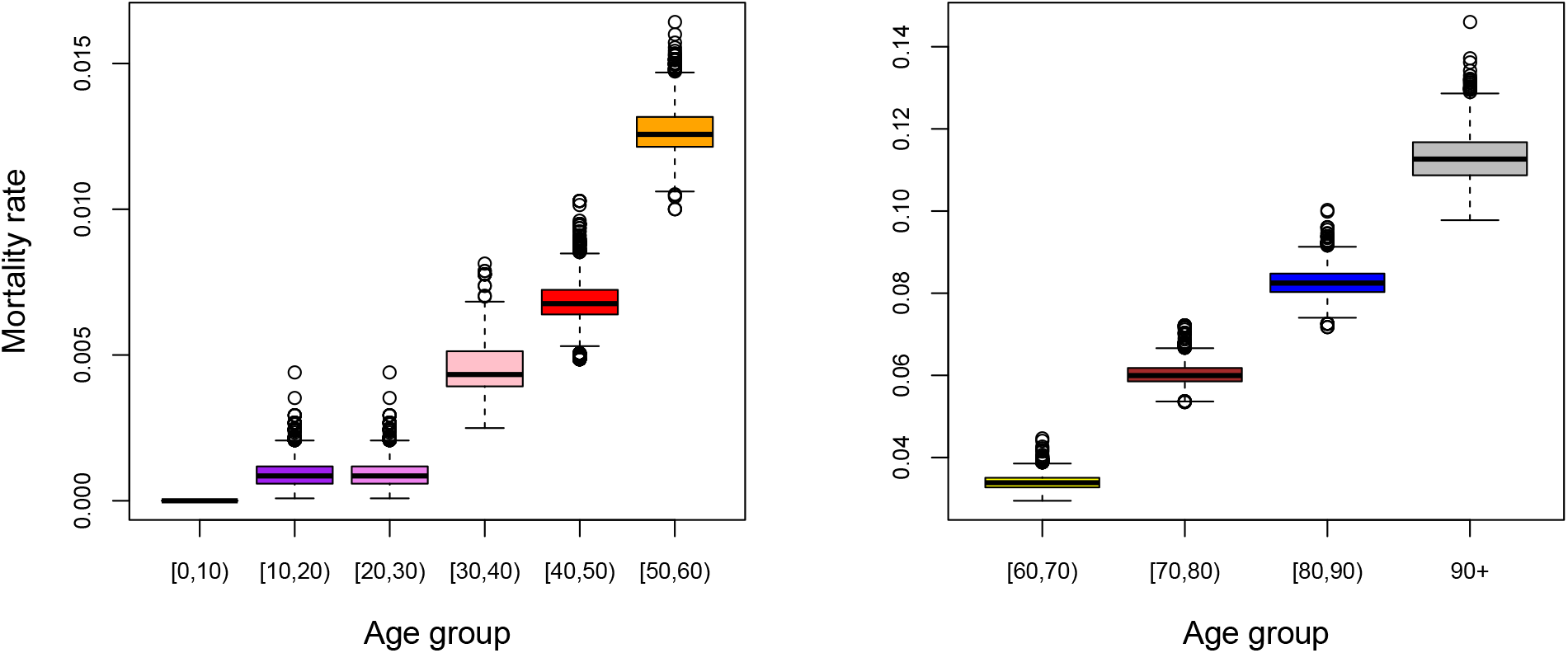
Boxplots of the marginal posterior distributions of the mortality rates by age group.

**Figure F5:**
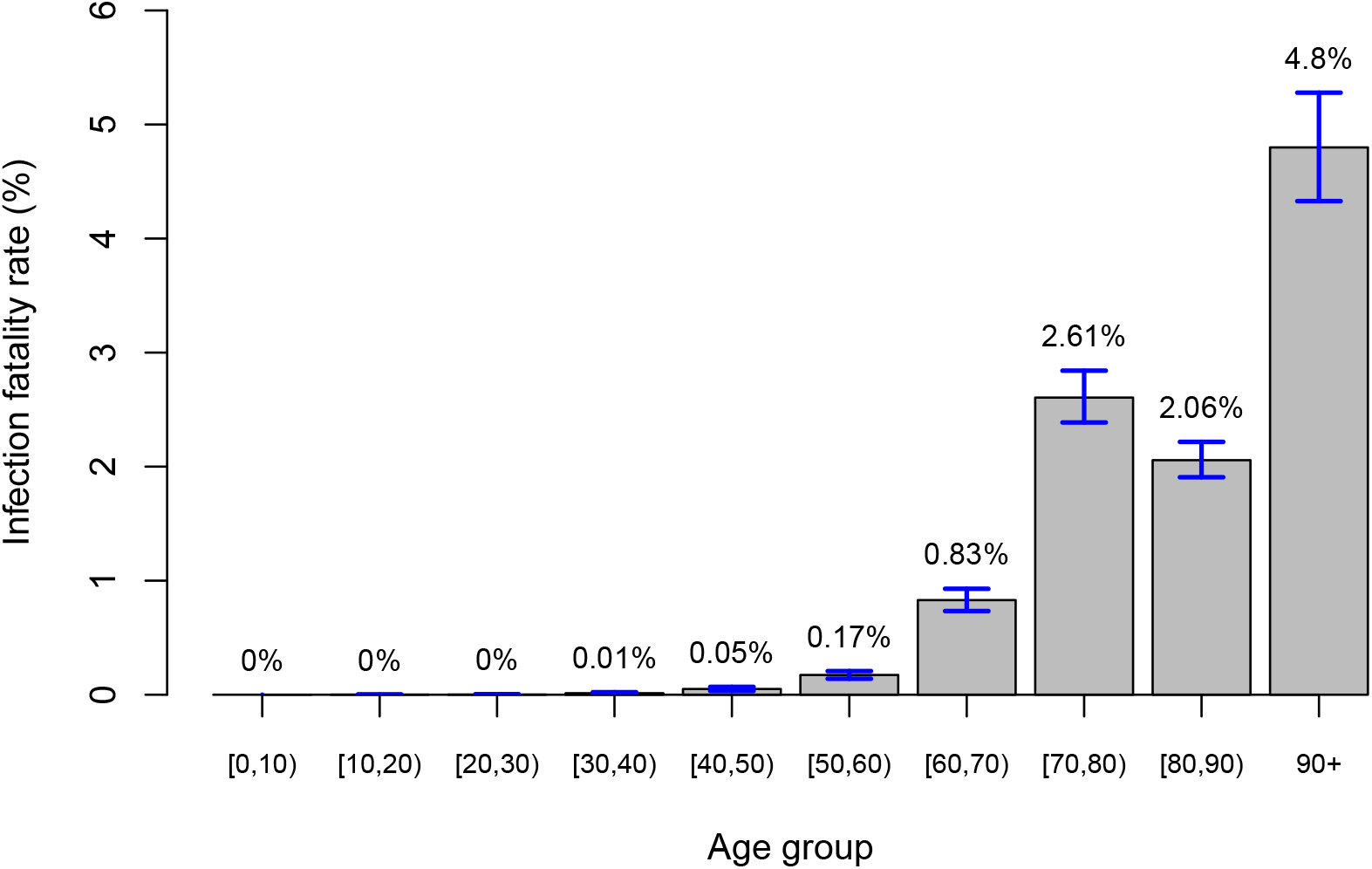
Infection fatality rates by age group with 95% credible intervals in blue.

### F.7 Cumulative number of hospitalizations (exit scenario analyses)

In Figure F6, we show the total number of hospitalizations over time in the different scenarios S7–S12. In general, the cumulative number of hospitalizations is highest in scenarios S10–S12 with the cumulative number of hospitalizations on average being comparable by the end of December, 2020 in scenarios S7–S9 on the one hand, and S10–S12 on the other hand.

**Figure F6:**
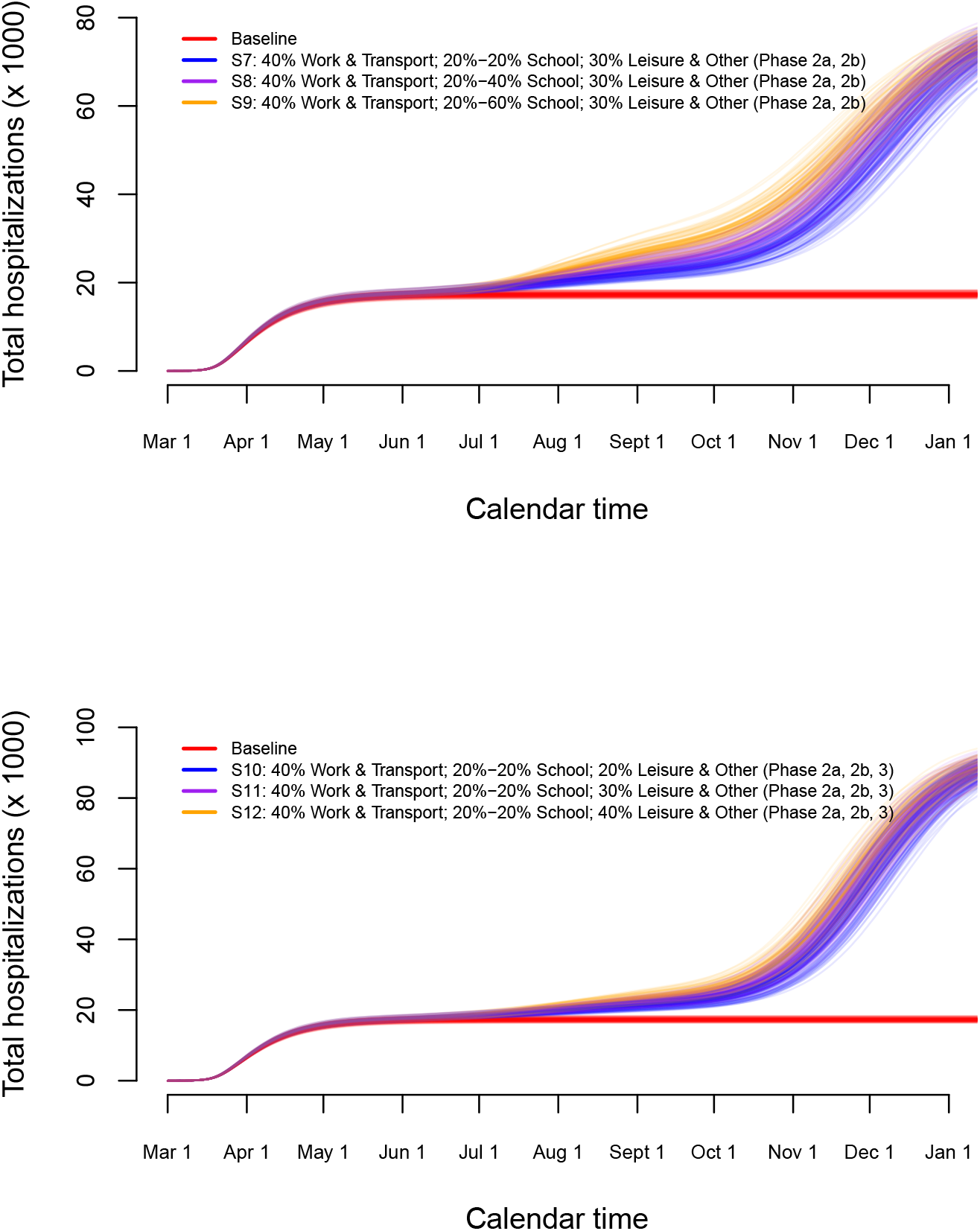
Long-term predictions of the impact of various exit strategies on the total number of hospitalizations over time.

### F.8 Sensitivity to transmission potential of children

As a sensitivity analysis, we studied the impact of varying the infectiousness of children on the final results. As mentioned in Section B.2, the role of children is still unclear albeit that some authors have tried to investigate the transmission potential of children. More specifically, children present a smaller viral load upon contracting the infection [1, 2, 3, 41, 42]. Furthermore, some authors claim that children have a reduced transmissibility [43, 44], albeit that it remains unclear whether this is due to a lower probability of presenting symptoms and differential transmissibility for asymptomatic versus symptomatic cases or directly by lowering infectiousness for both asymptomatic and symptomatic children.

We present stochastic simulation results based on a 50% reduction in the infectiousness of symptomatic and asymptomatic children in age category [0, 10). Posterior measures are very similar with only a small increase in the basic reproduction number *R*_0_ (3.021, 95% credible interval: 2.987, 3.056).

In Figures F7 and F8, we depict similar exit scenarios as the long-term scenarios S7–S12 presented in the main text. In general, the reduction in infectiousness leads to a decrease in peak size of the next wave of hospitalizations with a small delay in timing of the peak.

**Figure F7:**
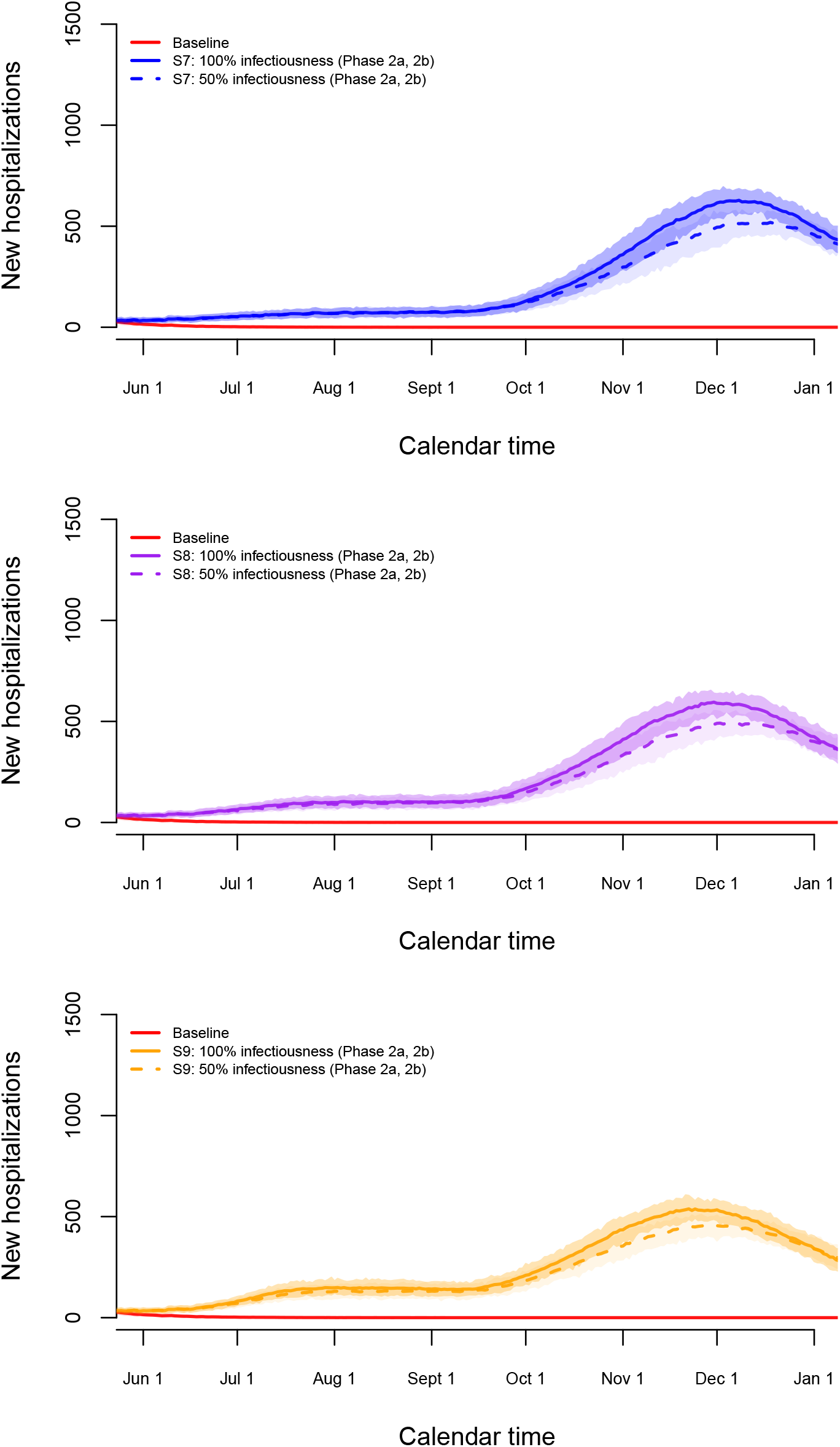
Long-term predictions of the impact of various exit strategies S7–S9 on the number of new hospitalizations.

**Figure F8:**
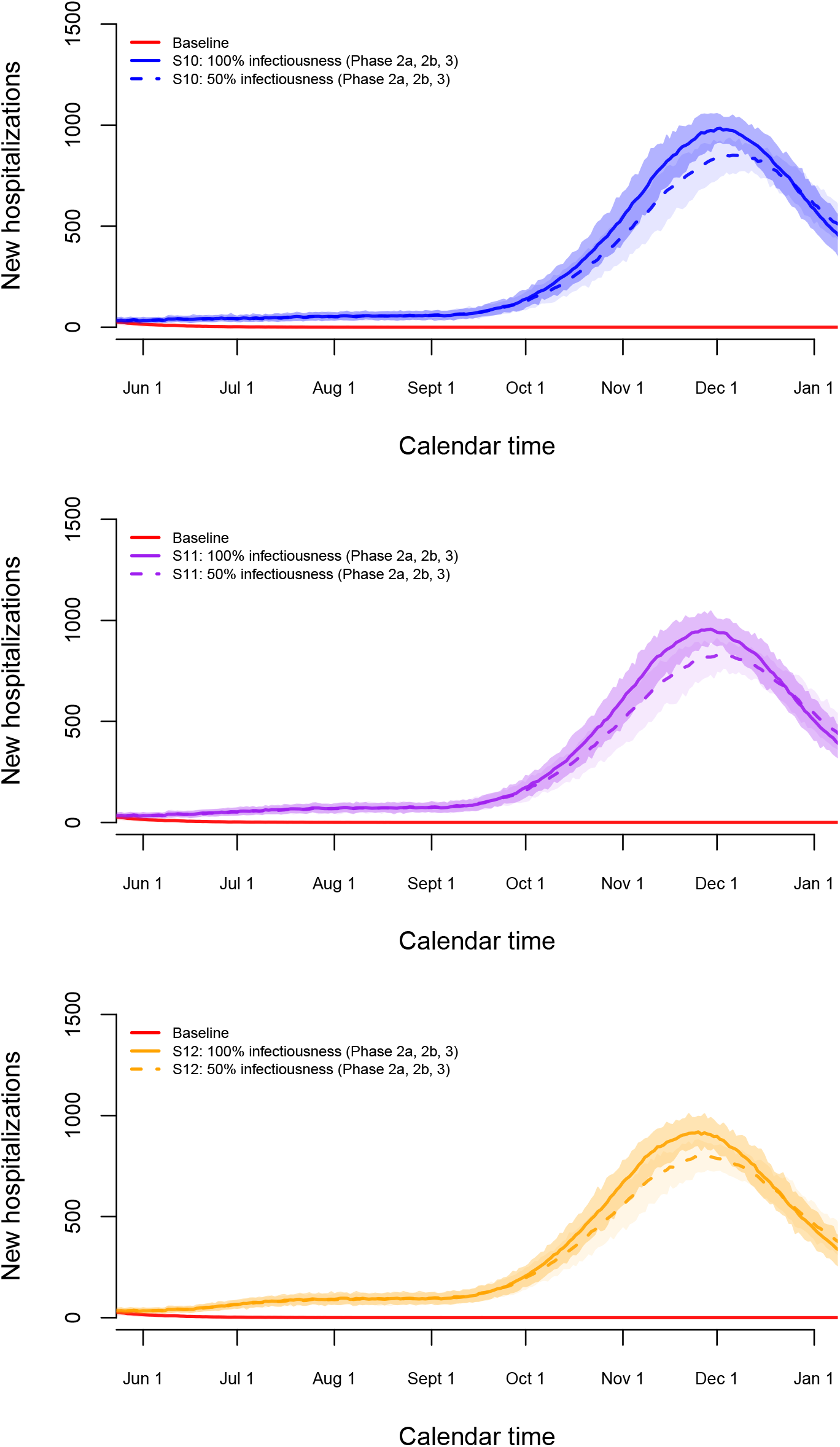
Long-term predictions of the impact of various exit strategies S10–S12 on the number of new hospitalizations.

### F.9 Sensitivity to diagnostic performance of serological IgG ELISA test

Changing the underlying sensitivity curve, leaving the presumed impact of intervention measures on the reduction of social contacts unchanged, mainly leads to a decrease in the posterior mean for *R*_0_ to 2.962 (95% credible interval (CI): 2.909, 3.018) and an increase for *ρ*^−1^, the average length of the latent period, with posterior mean 5.101 (95% CI: 5.016, 5.190). In Figure F9, the impact on the estimated overall prevalence over time is depicted. The estimated overall prevalence and corresponding 95% credible interval are shown for the original sensitivity curve (black dashed line with gray shaded area) and the alternative sensitivity curve (red solid line with red shaded area) as presented in Appendix E. In case of the different sensitivity curve the estimated prevalence is lower which is as expected given the faster detectability of IgG antibodies after SARS-CoV-2 infection. The estimated seroprevalence at the cross-sectional time points is almost identical for the two sensitivity curves (not shown).

**Figure F9:**
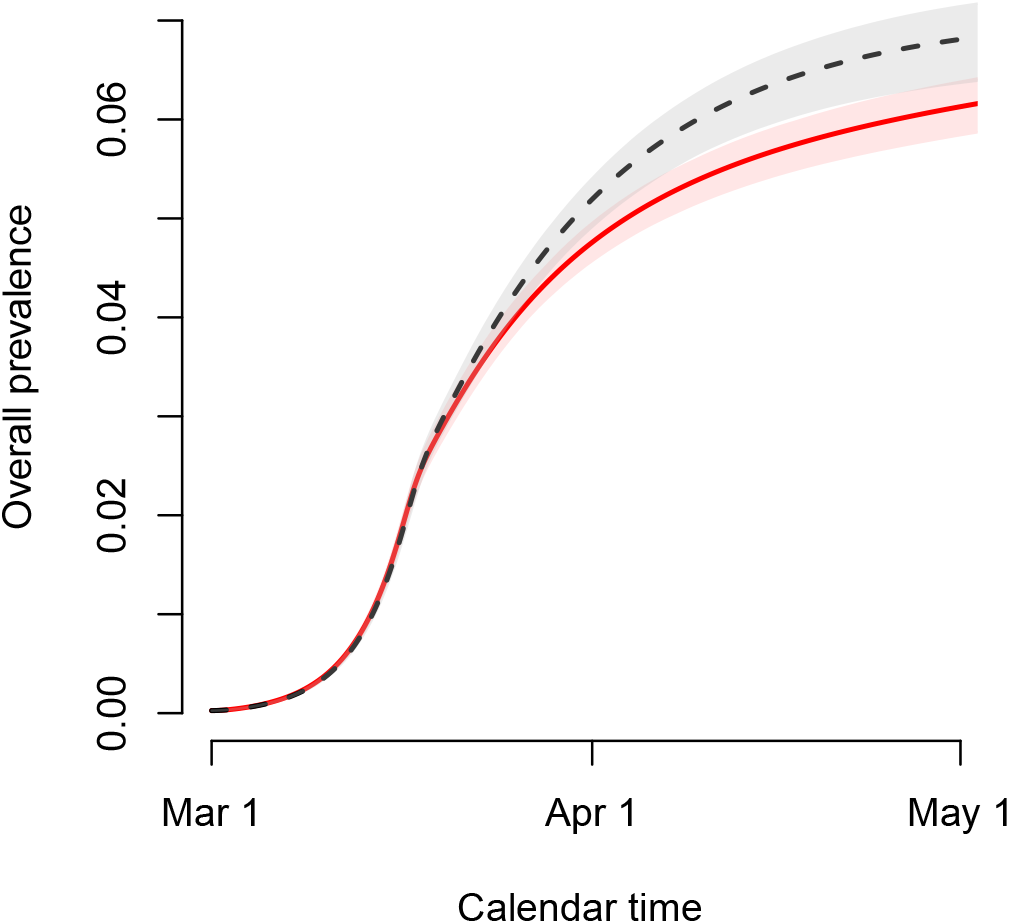
Estimated time-dependent prevalence of COVID-19 under different assumptions for the sensitivity of the IgG ELISA test.

In addition, we present the IFRs under different assumptions with regard to the sensitivity curve in Figure F10. A small increase in estimated average IFRs is observed for the higher age categories in case of the new sensitivity curve (red line in Figure E2).

**Figure F10:**
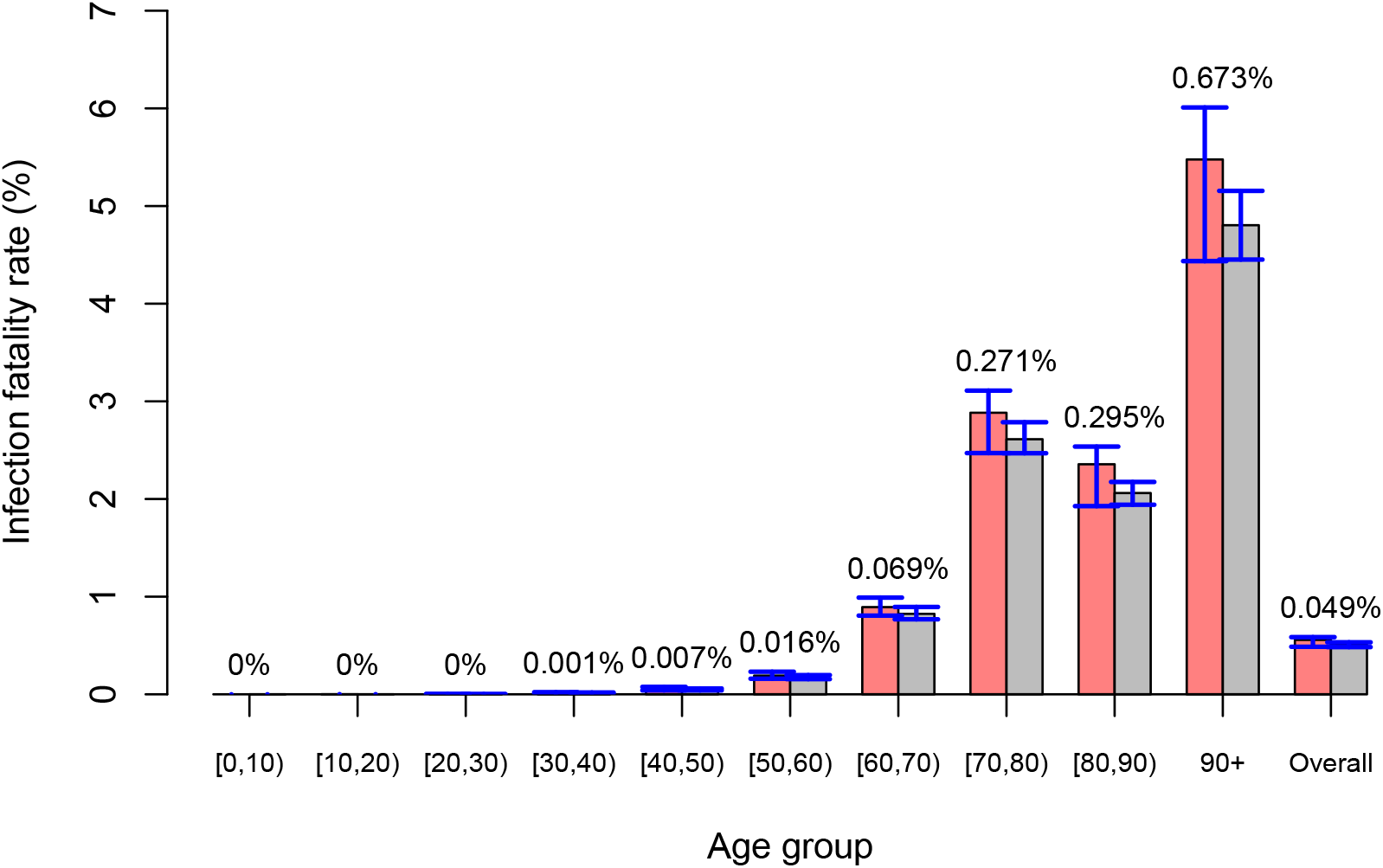
Infection fatality rates by age group with 95% credible intervals in blue and different sensitivity curves for the IgG ELISA test. Differences in mean IFRs between the two analyses indicated on top of the grouped bars per age category.

